# Plasma p-tau181/Aβ_1-42_ ratio predicts Aβ-PET status and correlates with CSF-p-tau181/Aβ_1-42_ and future cognitive decline

**DOI:** 10.1101/2022.03.13.22272320

**Authors:** Christopher Fowler, Erik Stoops, Stephanie Rainey-Smith, Eugeen Vanmechelen, Jeroen Vanbrabant, Nele Dewit, Kimberley Mauroo, Paul Maruff, Christopher C. Rowe, Jurgen Fripp, Qiao-Xin Li, Pierrick Bourgeat, Steven J. Collins, Ralph N. Martins, Colin L. Masters, James D. Doecke

## Abstract

**Background:** In Alzheimer’s disease, plasma Aβ_1-42_ and p-tau predict high amyloid status from Aβ-PET, however the extent to which combination of both plasma assays predict remains unknown.

**Methods:** Prototype Simoa assays were used to measure plasma samples from cognitively normal (CN) and symptomatic adults in the Australian Imaging, Biomarkers and Lifestyle (AIBL) study.

**Results:** The p-tau181/Aβ_1-42_ ratio showed the best prediction of Aβ-PET across all participants (AUC=0.905, 95%CI:0.86-0.95) and in CN (AUC=0.873; 0.80–0.94), and symptomatic (AUC=0.908; 0.82–1.00) adults. Plasma p-tau181/Aβ_1-42_ ratio correlated with CSF-p-tau181 (Elecsys®, Spearman’s ρ=0.74, P<0.0001) and predicted abnormal CSF Aβ (AUC=0.816, 0.74-0.89). The p-tau181/Aβ_1-42_ ratio also predicted future rates of cognitive decline assessed by AIBL PACC or CDR-SOB (P<0.0001).

**Discussion:** Plasma p-tau181/Aβ_1-42_ ratio predicted both Aβ-PET status and cognitive decline, demonstrating potential as both a diagnostic aid and as a screening and prognostic assay for preclinical Alzheimer’s disease trials.

## INTRODUCTION

Alzheimer’s disease (AD), the most common neurodegenerative dementia, is characterized by a preclinical and prodromal phase of >20 years, where amyloid beta (Aβ) accumulates as plaques in the brain extracellular environment, and with aggregation of tau in neurofibrillary tangles, drives neurodegeneration that gives rise to cognitive decline and ultimately dementia [1]. Aβ fragments exist in a number of distinct bioavailable pools, with the more insoluble load measurable by positron emission topography (Aβ-PET) and soluble Aβ detectable in the CSF [2-5]. Soluble phosphorylated tau (p-tau) can be detected in CSF, and PET can image insoluble aggregates of tau, all of which can aid AD diagnosis in research settings [6-8]. However, the use of PET and CSF to detect Aβ and tau in clinical or community settings is limited by the cost, requirements for specific expertise and equipment, and risk of adverse events. This restricts their use in programs seeking to identify individuals at risk for AD.

The advent of ultra-high sensitivity detection methods, including immunoprecipitation-mass spectrometry and single molecule array detection (including Simoa and other platforms) has enabled Aβ and p-tau species to be measured in blood plasma with increased dynamic range. When analyzed with specific detector and capture antibodies, plasma Aβ and p-tau levels are associated strongly with levels of the same biomarkers measured from CSF and with PET. For example plasma Aβ_1-42_ and the Aβ_1-42/1-40_ ratio predict brain Aβ burden, and tau measured at different phosphorylation sites (threonine_181_ (p-tau181), threonine_217_ (p-tau217), threonine_231_ (p-tau231)) shows concordance with CSF p-tau levels (concordance ∼0.8) and can predict Aβ-PET burden (AUC values ranging from 0.8 through 0.9) [9-14]. Clinical-pathological models show that understanding about the presence and severity of AD is improved when markers of amyloid and tau levels are considered simultaneously [15-18], however, to date, only one small study has investigated how combinations of plasma levels of Aβ and p-tau relate to Aβ-PET burden [19]. The first aim of this study was to investigate whether two different plasma p-tau markers (p-tau181 and p-tau231), incorporated into a ratio with Aβ, can improve predictions of Aβ burden over single analytes in comparison to PET or CSF sampling. Additionally, these analytes were investigated for an association with future cognitive decline. The second aim was to explore how ratios of plasma tau and amyloid were associated with disease progression.

## METHODS

### Study participants

The Australian Imaging, Biomarkers and Lifestyle (AIBL) study is a prospective longitudinal cohort of adults over the age of 60 designed to understand the natural history of AD, with recruitment and testing procedures described in detail [20]. Participants undergo 18 monthly neuropsychological and clinical assessment and blood donation, with AIBL clinical classification confirmed by an expert clinical panel consisting of a neurologist, geriatrician, and neuropsychologist; all blinded to biomarker status. All participants from the AIBL study, who had been classified clinically as being cognitively normal (CN), with mild cognitive impairment (MCI) or dementia, and who had both CSF samples and Aβ-PET scans were used in this study. Ethical approval was provided through St Vincent’s Health and Hollywood Private Hospital, and all participants provided written informed consent.

### Biospecimen collection

Blood collection was conducted on overnight fasting participants between 9:00am and 10:30am in K3-EDTA tubes (7.5 mL S-monovette 01.1605.008, Sarstedt, Australia) containing pre-added prostaglandin E1 (33 ng/mL of whole blood, Sapphire Biosciences, Australia) to prevent platelet activation, a potential source of peripheral Aβ. To generate plasma, blood was centrifuged at room temperature at 200g for 10 minutes to collect platelet-rich plasma, which was then spun at 800g for 10 minutes, aliquoted into 0.5 mL aliquots (2D cryobankIT, NUN374088), snap frozen within 2 hours of collection and then stored in vapor phase liquid nitrogen (LN_2_).

CSF was collected in the morning following overnight fasting via lumbar puncture using a Temena (Polymedic®, EU) spinal needle micro-tip (22/27G × 103 mm; CAT 21922-27). CSF was collected by aspiration or gravity drip into 15 mL polypropylene tubes (Greiner Bio-One188271) on wet ice, centrifuged within 1 hour at 2000g for 10 minutes at 4°C, transferred to a new 15 mL tube to remove any gradient effect and aliquoted into 2D NUNC cryovials and stored in vapor phase LN_2_.

Within AIBL, CSF collection is non-compulsory and can have intermittent follow up collections. In this paper Assessment 1 refers to the plasma matching the first CSF collection, and Assessment 2 refers to plasma collected matching a second CSF collection, either at 18 month or longer follow up intervals.

### Clinical

Calculation of the Alzheimer’s Disease Cooperative Study Preclinical Alzheimer Cognitive Composite in AIBL (AIBL-PACC) has been described [21]. For each assessment, an individual’s AIBL-PACC is computed by averaging the baseline-standardized scores of the Mini-Mental State Examination (MMSE), California Verbal Learning Test-II (CVLT-II), Logical Memory II, and Digit Symbol-Coding [22]. For analyses using both plasma biomarker and longitudinal cognition, the baseline clinical classification and cognitive status of individuals was aligned with the Assessment at which the plasma and CSF collection occurred first (Assessment 1). Longitudinal analyses then used all available cognitive measurements with respect to the plasma biomarker baseline (up to nine 18-month assessments). The number of Assessments available with cognitive data for participants in CN/CI groups are shown in Supplementary Table 1.

### Aβ-PET imaging

Aβ-PET imaging was performed with four different radiotracers: ^11^C-Pittsburgh compound B (PiB), ^18^F-NAV4694 (NAV), ^18^F-Flutemetamol (FLUTE) or ^18^F-Florbetapir (FBP). Aβ-PET scans were spatially normalized using CapAIBL [23] and the Centiloid (CL) method was applied [24, 25]. Aβ-PET (positive/negative) groups were derived using three CL thresholds: CL< 15, 20 and 25 as negative, ≥ 15, 20 and 25 as positive. Reporting on three thresholds has been used here, as variable CL thresholds have been utilized in the literature, with < 15 CL selected as a threshold for no neuritic plaques (CL threshold when calculated as +2 standard deviations from young normal ranging between 8-16 CL depending upon the tracer, and < 12 when compared to neuropathology), and ≥ 20 and ≥ 25 CL frequently used as the threshold for amyloid positivity corresponding to moderate plaque density [26-28]. Increasing the positivity threshold improves the correlation between neuropathological and clinicopathological diagnosis, but also allows for Consortium to Establish a Registry for Alzheimer Disease (CERAD) sparse neuritic plaques in a negative scan. CL data was not available for all participants at each of the assessments where plasma was collected. For Assessments 1 and 2, there were 169 and 65 participants from a total of 233 and 100 respectively with Aβ-PET measured at the same assessment as the plasma. For the remaining participants, the Aβ-PET status was imputed by binning the CL value (into bins derived via the 15, 20 and 25 CL thresholds) if a positive scan occurred prior to plasma collection and a negative scan post plasma collection.

### CSF analysis

CSF analysis in AIBL has been described [15] using the Roche Elecsys electrochemiluminescence immunoassays for Aβ_1-42_, Elecsys Total (t-)tau and Elecsys p-tau181, run on cobas e 601, cobas e 602 and MODULAR ANALYTICS E170 analyzers. A total of 155 participants had Elecsys measurements at the same time as the plasma (CN N = 106, MCI N = 28, AD N = 21). AIBL participant CSF and plasma samples were selected given biospecimen availability.

### Assays and Analytics

The set-up of the Amyblood (Simoa) assay was essentially as described in Thijssen et al, 2021 [29]. In short, C-terminal monoclonal antibodies (mAb), ADx102 and ADx103, are coupled to the paramagnetic carboxylated beads. mAb ADx101 was used as detection antibody in biotinylated form, with an antibody/biotin ratio of 32. Samples were diluted respectively 4 and 20 times for Aβ_1-42_ and Aβ_1-40_. Both assays were performed on the automated Quanterix Simoa HD-X platform using a 2-step protocol (80-7 cadences).

### Plasma p-tau assay

In the plasma p-tau protocol the phospho-specific mAbs ADx252 and ADx253 were conjugated to paramagnetic beads [30]. ADx252 is specific to p-tau181 and non-reactive towards p-threonine_175_ (p-tau175) nor p-tau231/p-serine_235_ (p-S235). ADx253 has a specificity towards p-tau231 and absence of reactivity towards p-S235 nor p-tau181/175. In addition, the phosphorylation of additional phospho-sites did not affect the reactivity (Supplementary Figures 1 and 2). Detection of p-tau was done using a N-terminal specific Tau mAb ADx204 in biotinylated form, that recognizes all tau forms except those phosphorylated at tyrosine_18_ [30] (ADx204 biotin/ratio of 32 for p-tau181 and ADx204/biotin ratio of 128 for p-tau231). After 8 minutes centrifugation at 10,000 g plasma samples were diluted 5-fold and were run on the Quanterix Simoa HD-X platform using a 2-step protocol of 80-14 cadences for the p-tau181 assay and 80-7 cadences for p-tau231. Calibration of both p-tau assays was done with a single synthetic peptide covering the relevant antibody epitopes. The seven calibrator points ranged between 50 pg/mL and 0.78 pg/mL, and 50 pg/mL and 0.39 pg/mL respectively, for p-tau181 and p-tau231. A five parameter curve fit algorithm with 1/Y^2^ weighting was used to convert AEB into p-tau concentrations. Details of the assay specifications are described in Supplementary Table 2.

P-tau assay phospho-specificity was assessed by checking sandwich assay reactivity using synthetic peptides containing the (non-)phosphorylated epitopes for each respective phospho-site threonine_181_ and/or threonine_231_ up to a concentration of 1000 pg/mL. Reactivity dependency of phosphorylation at other phospho-sites was also checked by using peptides with non-phosphorylation on threonine_175_ and serine_235_ up to a concentration of 1000 pg/mL (Supplementary Figures 1 and 2).

Further information on sample analysis, QC panel testing, run to run variability, accuracy and intra-run precision are in the Supplementary Materials.

### Statistical analyses

Statistical analyses were performed to test four main hypotheses; 1) determine which of the biomarkers and their ratios have the largest mean difference between Aβ-PET groups, 2) investigate the capability of each of the biomarkers and their ratios to predict Aβ-PET groups, 3) investigate how well plasma biomarkers correlate with their corresponding CSF biomarkers, and compare how well plasma biomarkers predict CSF Aβ_1-42_ compared with predicting Aβ-PET, and 4) using the best performing plasma biomarker, assess its ability to predict cognitive decline using both a late and early parameter of cognitive change (linear mixed effects models). Full details for the statistical analyses are in the Supplementary Materials. Only tables/plots for Assessment 1 are shown in the main text, while results from Assessment 2 are shown in Supplementary Materials.

## RESULTS

### Study demographic characteristics

Of the 233 participants with plasma collected at their first assessment, and the 100 participants with plasma taken at their second, 45% in Assessment 1, and 42% at Assessment 2 were Aβ-PET^+^. There was no difference in age or gender between the Aβ-PET groups (P>0.05), however Aβ-PET^+^ participants were more likely to carry at least one *APOE ε*4 allele (P<0.0001), have higher CL values (P<0.0001) and perform worse on MMSE, the AIBL PACC score and CDR-SOB (P<0.0001). Comparisons of sample demographics for Assessment 1 are shown in Table 1, and for Assessment 2 in Supplementary Table 3. Comparison of group mean biomarkers levels showed large differences between Aβ-PET groups for the p-tau181 (Cohen’s D: 1.19) and p-tau181/Aβ_1-42_ ratio (Cohen’s D: 1.29) at both Assessments (P<0.0001, Supplementary Table 4, Figure 1).

**Figure 1:**
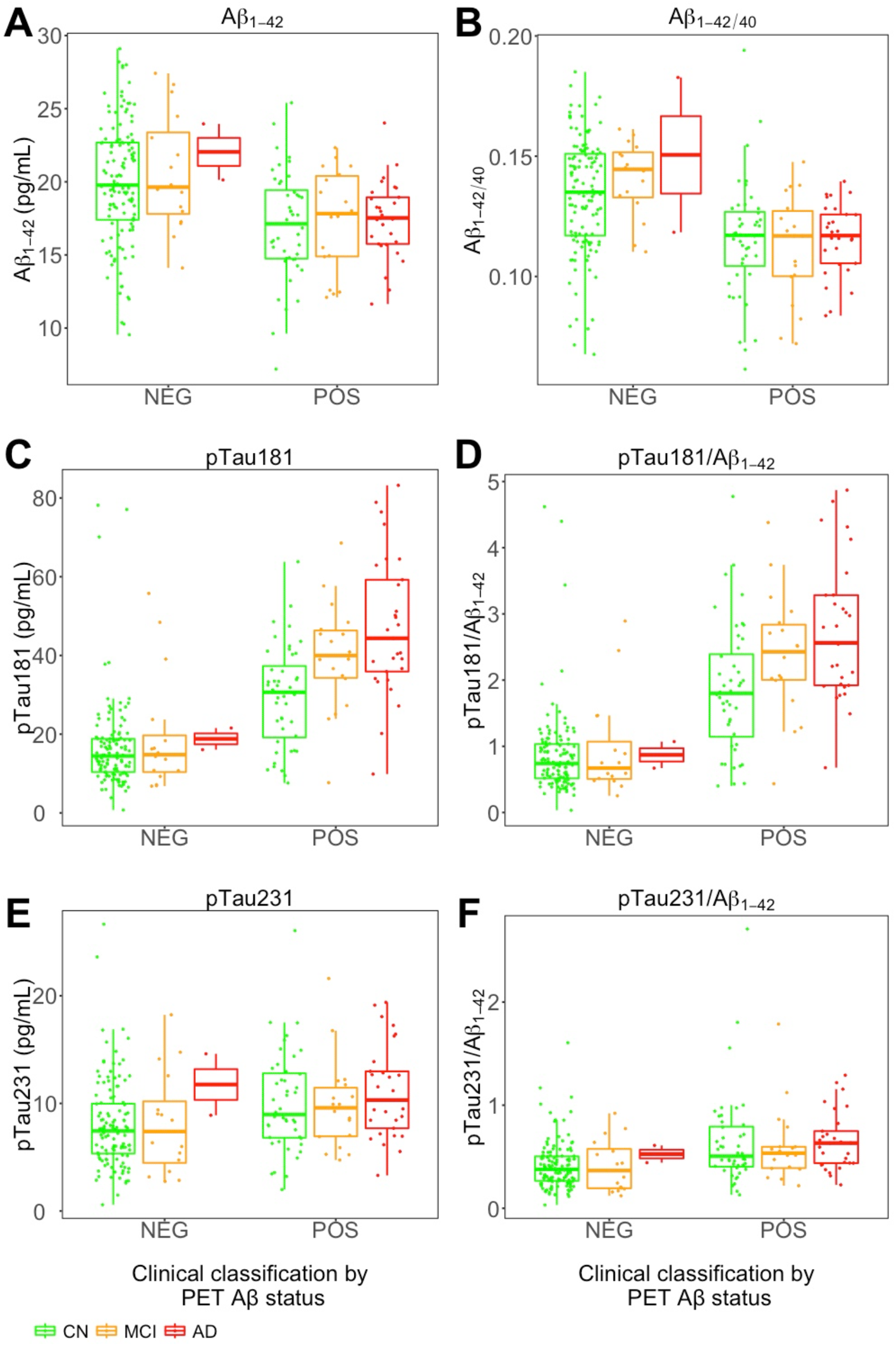
Box and whisker plot for Aβ_1-42_, p-tau181 & p-tau231, Aβ_1-42/1-40_ ratio, p-tau181/Aβ_1-42_ ratio & p-tau231/Aβ_1-42_ ratio at Assessment 1. Individual markers (Aβ_1-40_, Aβ_1-42_ p-tau181 & p-tau231) are measured in pg/mL. A) Aβ_1-42_, B) Aβ_1-42/40_, C) pTau181, D) pTau181/Aβ_1-42,_ E) pTau231, F) pTau231/Aβ_1-42_. Plots represent the 5 number summary, with the median, first and third quartiles shown in each box.

**Table 1:** Study demographic characteristics.

| | Total Sample | Aβ<sup>-</sup> | Aβ<sup>+</sup> | p-value |
| --- | --- | --- | --- | --- |
| N (%) | 233 | 142 (61%) | 91 (39%) |  |
| Gender Male, N (%) | 124 (53%) | 72 (51%) | 52 (57%) | 0.34 |
| Mean Age, years (SD) | 72.9 (6.1) | 72.4 (6.3) | 73.5 (5.8) | 0.18 |
| APOE ε4 Carriage, N (%) | 74 (32%) | 26 (18%) | 48 (53%) | <0.0001 |
| Tracer Florbetapir N % | 60 (26%) | 44 (26%) | 16 (26%) |  |
| Tracer Flutemetamol N % | 73 (31%) | 42 (31%) | 31 (31%) |  |
| Tracer NAV/PiB N % | 100 (43%) | 56 (43%) | 44 (43%) | 0.069 |
| Mean CL (SD) | 34.2 (45.2) | 1.8 (9.6) | 80 (34.9) | <0.0001 |
| Diagnosis CN N % | 168 (72%) | 124 (53%) | 44 (19%) |  |
| Diagnosis MCI N % | 34 (15%) | 16 (7%) | 18 (8%) |  |
| Diagnosis AD N % | 31 (13%) | 2 (1%) | 29 (12%) | <0.0001 |
| Median MMSE, (MAD) | 28 (1.5) | 29 (1.5) | 28 (3) | <0.0001 |
| Median CDR SB, (MAD) | 0 (0) | 0 (0) | 0.5 (0.7) | <0.0001 |
| Mean AIBL PACC (SD) | 233 | 142 (61%) | 91 (39%) |  |

Characteristics measured at visit 1 and compared between Aβ-PET groups using a CL threshold at 20CL.

### Plasma biomarkers to predict Aβ-PET

Receiver operating characteristic (ROC) analyses of plasma data to predict Aβ-PET groups at both assessments and across three CL thresholds demonstrated strong predictive capability for both the p-tau181 marker alone and its ratio with Aβ_1-42_ (Table 2, Supplementary Tables 5, 6 & 7, Supplementary Figure 1). Using the ratio to predict Aβ-PET groups (CL threshold at 20CL) performed significantly better than both the p-tau181 marker alone and any combination of markers as tested via multivariate modelling. p-tau231 alone did not significantly differ from the base model and further information on the predictive capability comparisons is presented in Supplementary Table 9. Concentrating on the p-tau181/Aβ_1-42_ ratio and restricting the sample set to the CN adults only (Assessment 1), predictive performance was reduced slightly (AUC complete group 0.88, CN 0.83), while in the MCI subgroup it increased to 0.91. Predictive performance was similar at Assessment 2, with higher AUC values despite the smaller sample size (Supplementary Table 5). Assessing the predictive capability in models using confounders age, gender *APOE ε*4 allele status and tracer showed only small improvements in AUC values (AUC [95%CI] unadjusted: 0.883 [0.83-0.93)], adjusted: 0.889 [0.84-0.94]). All models including plasma p-tau181 out-performed the base model (P<0.0001, Supplementary Table 7, Supplementary Figure 2 & Supplementary Figure 3). Positive and negative predictive values for the p-tau181/Aβ_1-42_ ratio remained similar whether using ROC models from the individual biomarkers, or from models including both biomarker and confounders age, gender tracer and *APOE ε*4 allele status at both assessments (Supplementary Table 8).

**Table 2:** AUC values across complete and stratified groups using CL threshold at 20

|  | Complete sample | CN | CI |
| --- | --- | --- | --- |
| Biomarker | AUC (95%CI) | AUC (95%CI) | AUC (95%CI) |
| N ($A\beta^-/A\beta^+$) | 142/91 | 124/44 | 18/47 |
| $A\beta_{1-42}$ | 0.705 (0.64 - 0.77) | 0.705 (0.62 - 0.79) | 0.741 (0.6 - 0.88) |
| $A\beta_{1-42/1-40}$ | 0.731 (0.69 - 0.81) | 0.728 (0.64 - 0.81) | 0.852 (0.74 - 0.96) |
| p-tau181 | 0.862 (0.81 - 0.92) | 0.808 (0.72 - 0.89) | 0.868 (0.75 - 0.98) |
| p-tau231 | 0.66 (0.59 - 0.73) | 0.646 (0.55 - 0.74) | 0.636 (0.47 - 0.8) |
| p-tau181/$A\beta_{1-42}$ | 0.883 (0.83 - 0.93) | 0.832 (0.75 - 0.91) | 0.908 (0.82 - 1) |
| p-tau231/$A\beta_{1-42}$ | 0.713 (0.65 - 0.78) | 0.692 (0.6 - 0.78) | 0.696 (0.54 - 0.85) |

### Biomarker correlation and agreement between plasma, CSF and PET

Associations between markers of tau and amyloid measured from plasma and CSF (Figure 2) were strongest for the p-tau181/Aβ_1-42_ ratio, with a Spearman’s Rho value of 0.75 (P<0.0001). p-tau181 and the Aβ_1-42/1-40_ ratio were the next strongest with Rho values of 0.53 and 0.45 (P<0.0001), whilst Aβ_1-42_ showed more variation between plasma and CSF Rho = 0.32 (P<0.0001). Assessing agreement between the plotted CL values and p-tau181/Aβ_1-42_ ratio values binned into quadrants, (Figure 3 & Supplementary Figure 2) demonstrated strong agreement between the two markers. Of the 99 participants at Assessment 1 with a CL value less than 20, 92 participants (93%) were both Aβ-PET^-^ and p-tau181/Aβ_1-42_ negative, whilst only 7 participants (7%) were Aβ-PET^-^ and p-tau181/Aβ_1-42_ positive. Of the 70 participants who were Aβ-PET^+^, 57 participants (81%) were also p-tau181/Aβ_1-42_ positive, whilst 13 participants (19%) were Aβ-PET^+^ and p-tau181/Aβ_1-42_ negative.

**Figure 2:**
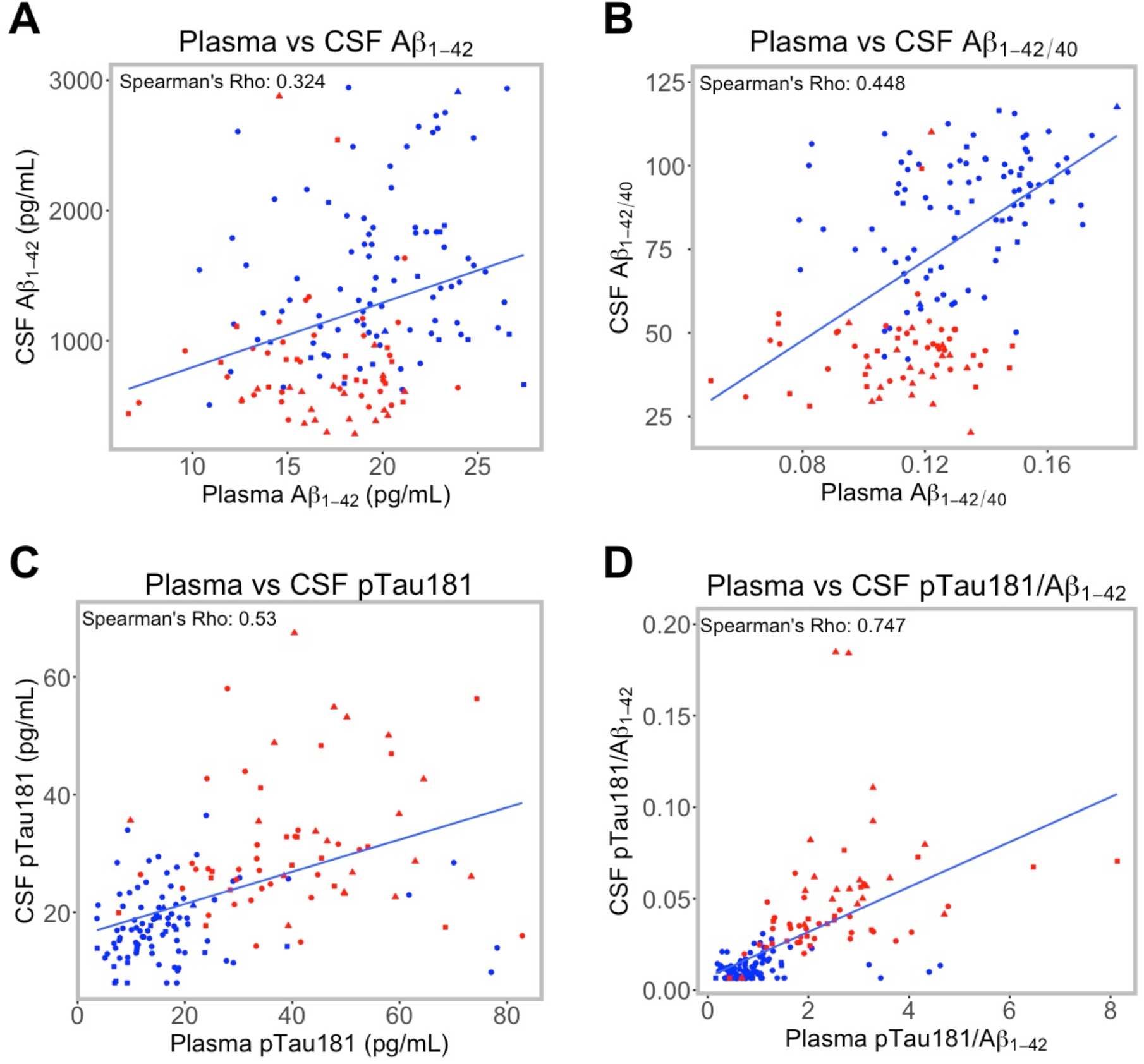
Correlation between plasma and CSF biomarkers. Sample size for correlation plots at visit one was N=155. Linear fit lines are drawn irrespective of Aβ-PET status. Red points represent participants who were Aβ-PET^+^; blue points represent participants who were Aβ-PET^-^. Circle points represent those participants who were CN, square points represent those participants with MCI, triangle points represent those participants with AD. A) Plasma vs CSF Aβ_1-42_, B) Plasma vs CSF Aβ_1-42/1-40_, C) Plasma vs CSF pTau181, D) Plasma vs CSF pTau181/Aβ_1-42_.

**Figure 3:**
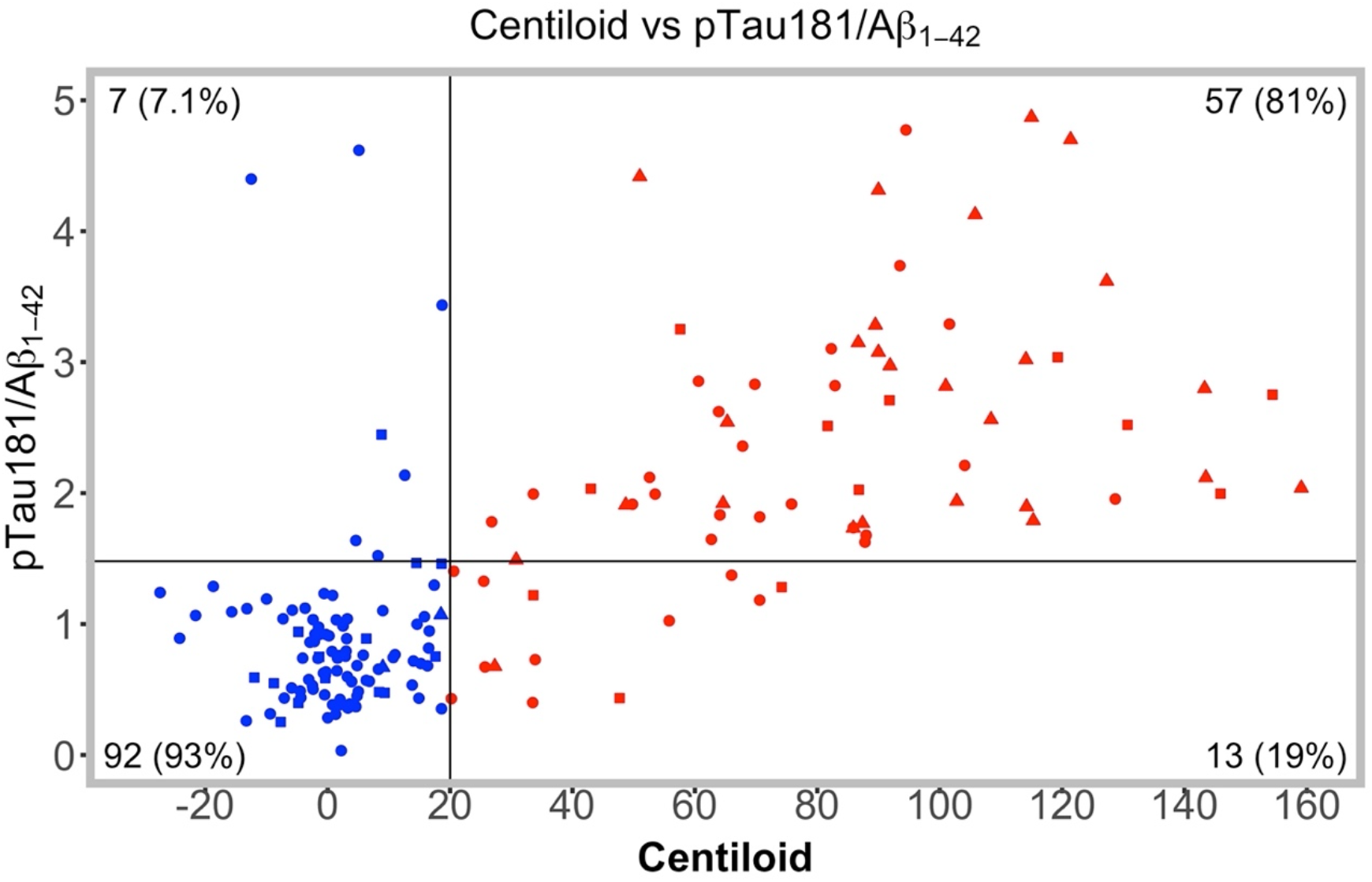
Agreement between Aβ-PET and plasma p-tau181/Aβ_1-42_ ratio at visit one. Threshold line for Aβ-PET was set at 20CL. Threshold for the p-tTau181/Aβ_1-42_ ratio was set using the Youden’s Index (1.48) from the ROC model for Aβ-PET status with the CL threshold set at 20CL at visit one. Red points represent participants who were Aβ-PET^+^; blue points represent participants who were Aβ-PET^-^. Circle points represent those participants who were CN, square points represent those participants with MCI, triangle points represent those participants with AD.

### Plasma vs CSF biomarkers to predict Aβ-PET status

Comparing the performance of Aβ_1-42_, p-tau181 and p-tau181/Aβ_1-42_ to predict the Aβ-PET status at each of the three thresholds (Supplementary Figure 3) in plasma (measured using Simoa assays) and CSF (measured using Elecsys assays), it was clear that the Aβ_1-42_ assay for CSF outperformed the plasma assay (@CL15 P=0.012, @CL20 P=0.0003, @CL25 P=0.0003), whilst the CSF p-tau181 assay performed no differently to the plasma assay (@CL15 P=0.887, @CL20 P=0.822, @CL25 P=0.856). For the ratio however there was no difference in AUC values between the plasma and CSF assay at 15CL (P=0.409), with weak but significant differences using the CL threshold at 20 (P=0.033) and at 25 (P=0.051). As for Aβ-PET, ROC analyses were performed using CSF Aβ_1-42_ as the outcome variable (Supplementary Table 9 & Supplementary Figure 1E & 1F). AUC values were lower for plasma biomarkers to predict CSF-Aβ as compared with predicting Aβ-PET status, however the p-tau181/Aβ_1-42_ ratio still had the highest AUC (0.82 [95%CI: 0.71 - 0.89]).

### Plasma p-tau181/Aβ_1-42_ predicts change in cognition at both early and late stages of AD

Analyses of associations between levels of the plasma p-tau181/Aβ_1-42_ ratio with change in cognition over time and adjusting for age, gender, tracer and *APOE ε*4 allele status showed a significant increase in CDR SoB (Figure 4D, P<0.0001) in the CI group, a significant increase in CDR SoB (Figure 4C, P=0.015) in the CN group and a significant decrease in the AIBL PACC score in the CN group over time (Figure 4A, P=0.0002). Repeating these analyses using the CSF p-tau181/Aβ_1-42_ ratio showed only the CDR SoB to have a significant difference in the change in cognition for those with a higher p-tau181/Aβ_1-42_ ratio as compared with a low p-tau181/Aβ_1-42_ ratio in the CI group (Supplementary Figure 4D, p=0.003).

**Figure 4:**
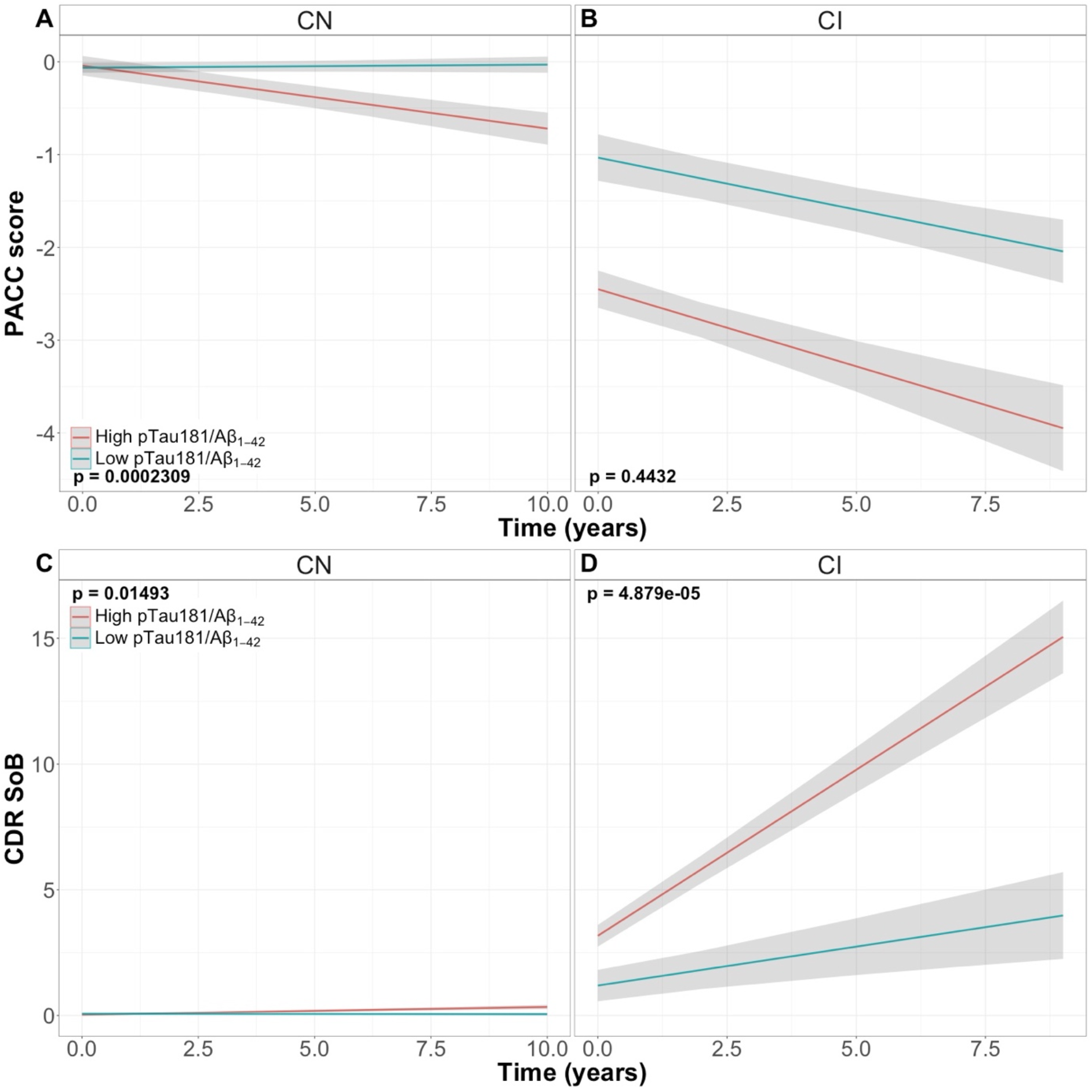
Cognitive decline in CDR SoB and the AIBL PACC score using the plasma between CN/CI groups and the pTau181/Aβ_1-42_ ratio as measured by linear mixed effects models. Time on the x-axis refers to the first AIBL assessment whereby the plasma was collected. Sample sizes for cognitive collection points for the plasma sample set are shown in Supplementary Table 1. The binary p-tau181/Aβ_1-42_ ratio was created using the Youden’s Index (1.48) created from the ROC model using plasma p-tau181/Aβ_1-42_ vs Aβ-PET using a CL threshold at 20CL.

## DISCUSSION

In the current study we have used the Amyblood assay on the ultra-sensitive Simoa platform to measure, in the AIBL cohort, plasma Aβ_1-40_ and Aβ_1-42_, and a prototype assay for measuring p-tau181 and p-tau231, to determine that p-tau181/Aβ_1-42_ ratio is the highest performing test compared to individual measurements in predicting amyloid positive status. Moreover, we observed that only the plasma p-tau181/Aβ_1-42_ ratio was correlated strongly with the CSF p-tau181/Aβ_1-42_ ratio, and that this plasma ratio predicted future cognitive decline within the CN sub group for PACC, and the CI sub group for CDR-SOB.

In a cohort of 233 participants, comprising CN, MCI and AD participants, the p-tau181/Aβ_1-42_ ratio yielded an AUC of 0.91 for determining amyloid positivity when analysed against a PET amyloid threshold of 25 CL. When separated into CN and MCI cohorts, the AUCs remained very high (CN =0.87, MCI= 0.91). The AUCs improved as the CL threshold was increased. This compares favourably with other Simoa Aβ and pTau analyses whose AUCs range between 0.66 to 0.88 [11, 31-33], and is similar to AUCs for the only current FDA-approved plasma test for determining amyloid positivity, which in a cohort of predominately CDR-SOB 0 had an AUC of 0.88 and 0.94 when run without and with other covariates of age and *APOE ε*4 on a mass spectrometry platform [34]. In the AIBL study cohort, addition of *APOE ε*4, sex, age and PET tracer only marginally improved the performance of the plasma test, potentially reducing the need for measuring other parameters when used as a trial-screening tool. Analyses incorporating p-tau181 improved significantly, although mildly, to the addition of covariates, possibly related to the age-dependent accumulation that CSF tau is known to display [35]. To our knowledge, only one other study investigated the ratio of p-tau181/Aβ_1-42_ in plasma in relation to Aβ-PET which produced an AUC of 0.89 in both CN and MCI participants [19]. The current work, conducted in in a larger cohort and in a different ethnic group, supports the utility of this measurement approach. Other works, while not using a ratio, have also identified that combination analyses on p-tau, Aβ and clinical features can better distinguish AD from unimpaired populations [36] than analyses of analytes alone.

Compared to CSF measures of p-tau181, Aβ_1-40_ and Aβ_1-42_, measured in a subset of this cohort on the Elecsys immunoassay, the ratio of p-tau181/Aβ_1-42_ in both the CSF and plasma was the only measure that was highly correlated, although measured on different platforms. The p-tau181/Aβ_1-42_ ratio has long been considered to have the best statistical power of the CSF markers for representing AD, as it incorporates both hallmarks of the AD disease pathway and represents a later stage of the disease pathway. Indeed this ratio in CSF has helped identify late stage changes in synaptic and neuronal degradation markers and inflammatory markers associated with AD [37], as well as show high concordance with Aβ-PET [15-17, 20]. Here analyses of the plasma markers at a second time point in this study showed increased performance of all markers, including the p-tau181/Aβ_1-42_ ratio, at predicting amyloid status at Assessment 2, which may reflect further accumulation of positive biomarker signals compared to a static baseline in the amyloid negative cohort. More analyses in other cohorts are needed to test if this plasma ratio can act as a proxy for the CSF measurements to suitably correlate with synaptic degradation biomarkers.

Determination of amyloid chronicity is an estimation of where an individual’s current level of amyloid sits temporally on an amyloid accumulation pathway. This accumulation pathway, which is non-linear in accumulation over the initial 20 year preclinical phase, and is independent of the individual’s starting age, requires biomarkers that can detect concurrent pathological hallmarks, including accumulation of p-tau, to aid understanding of the near term risk of phenotypic cognitive decline. Analyses here into longitudinal cognitive outcomes with up to 10 years follow-up, showed that the ratio was able to identify those with early cognitive change as demonstrated via the AIBL PACC, and showed a clear separation in the CDR-SOB in both CN and CI groups. The ability to detect cognitive change in those classed as CN further demonstrates the ratio’s capacity to detect the minority of this population with Amyloid, thus acting as an alternative to PET imaging. For CDR-SOB, those with the high ratio performed significantly worse compared with those with a low ratio, suggesting that even once impaired, participants cognitive performance can be staged as per their ratio level; a very useful trait for clinical trial recruitment. These data suggest that the ratio may assist in the determination of disease chronicity and should be investigated across more longitudinal plasma collection points.

The p-tau181 analyte alone was the next best performing measurement, while the p-tau231 analyte did not perform better than the Aβ measurements, nor add anything when in ratio with the Aβ for correlating with Aβ-PET or CSF-Aβ levels. Further longitudinal analysis, and incorporation of Tau-PET measurements into the analysis may reveal temporal specificities for changes in p-tau231 more subtle than what can be detected in this work.

Presently the FDA has provisionally approved the first immunogenic anti-amyloid therapy for mild AD and there is a need for cheap, non-invasive blood tests to not only predict amyloid burden to assist in identifying participants at risk for developing AD, and to confirm AD diagnosis, but to aid in determining the temporal location along the biomarker accumulation pathway. The plasma p-tau181/Aβ_1-42_ ratio predicts here Aβ-PET status but also hints at predicting near term future cognitive decline more accurately than Aβ-PET status alone.

## Data Availability

Most data produced in the present study are available upon reasonable request to the authors

## References

[1] Busche MA, Hyman BT. Synergy between amyloid-beta and tau in Alzheimer’s disease. Nat Neurosci. 2020;23:1183–93.

[2] Rowe CC, Pejoska S, Mulligan RS, Jones G, Chan JG, Svensson S, et al. Head-to-head comparison of 11C-PiB and 18F-AZD4694 (NAV4694) for beta-amyloid imaging in aging and dementia. J Nucl Med. 2013;54:880–6.

[3] Roberts BR, Lind M, Wagen AZ, Rembach A, Frugier T, Li QX, et al. Biochemically-defined pools of amyloid-beta in sporadic Alzheimer’s disease: correlation with amyloid PET. Brain. 2017;140:1486–98.

[4] Salloway S, Gamez JE, Singh U, Sadowsky CH, Villena T, Sabbagh MN, et al. Performance of [(18)F]flutemetamol amyloid imaging against the neuritic plaque component of CERAD and the current (2012) NIA-AA recommendations for the neuropathologic diagnosis of Alzheimer’s disease. Alzheimers Dement (Amst). 2017;9:25–34.

[5] Jack CR, Jr., Bennett DA, Blennow K, Carrillo MC, Feldman HH, Frisoni GB, et al. A/T/N: An unbiased descriptive classification scheme for Alzheimer disease biomarkers. Neurology. 2016;87:539–47.

[6] Villemagne VL, Fodero-Tavoletti MT, Masters CL, Rowe CC. Tau imaging: early progress and future directions. Lancet Neurol. 2015;14:114–24.

[7] Jack CR, Jr., Bennett DA, Blennow K, Carrillo MC, Dunn B, Haeberlein SB, et al. NIA-AA Research Framework: Toward a biological definition of Alzheimer’s disease. Alzheimers Dement. 2018;14:535–62.

[8] Pascoal TA, Shin M, Kang MS, Chamoun M, Chartrand D, Mathotaarachchi S, et al. In vivo quantification of neurofibrillary tangles with [(18)F]MK-6240. Alzheimers Res Ther. 2018;10:74.

[9] Nakamura A, Kaneko N, Villemagne VL, Kato T, Doecke J, Doré V, et al. High performance plasma amyloid-β biomarkers for Alzheimer’s disease. Nature. 2018;554:249–54.

[10] Barthelemy NR, Horie K, Sato C, Bateman RJ. Blood plasma phosphorylated-tau isoforms track CNS change in Alzheimer’s disease. J Exp Med. 2020;217.

[11] Karikari TK, Pascoal TA, Ashton NJ, Janelidze S, Benedet AL, Rodriguez JL, et al. Blood phosphorylated tau 181 as a biomarker for Alzheimer’s disease: a diagnostic performance and prediction modelling study using data from four prospective cohorts. Lancet Neurol. 2020;19:422–33.

[12] Thijssen EH, La Joie R, Wolf A, Strom A, Wang P, Iaccarino L, et al. Diagnostic value of plasma phosphorylated tau181 in Alzheimer’s disease and frontotemporal lobar degeneration. Nat Med. 2020;26:387–97.

[13] Ashton NJ, Pascoal TA, Karikari TK, Benedet AL, Lantero-Rodriguez J, Brinkmalm G, et al. Plasma p-tau231: a new biomarker for incipient Alzheimer’s disease pathology. Acta Neuropathol. 2021;141:709–24.

[14] Ossenkoppele R, Reimand J, Smith R, Leuzy A, Strandberg O, Palmqvist S, et al. Tau PET correlates with different Alzheimer’s disease-related features compared to CSF and plasma p-tau biomarkers. EMBO Mol Med. 2021;13:e14398.

[15] Doecke JD, Ward L, Burnham SC, Villemagne VL, Li QX, Collins S, et al. Elecsys CSF biomarker immunoassays demonstrate concordance with amyloid-PET imaging. Alzheimers Res Ther. 2020;12:36.

[16] Campbell MR, Ashrafzadeh-Kian S, Petersen RC, Mielke MM, Syrjanen JA, van Harten AC, et al. P-tau/Abeta42 and Abeta42/40 ratios in CSF are equally predictive of amyloid PET status. Alzheimers Dement (Amst). 2021;13:e12190.

[17] van Harten AC, Wiste HJ, Weigand SD, Mielke MM, Kremers WK, Eichenlaub U, et al. Detection of Alzheimer’s disease amyloid beta 1-42, p-tau, and t-tau assays. Alzheimers Dement. 2021.

[18] Willemse EAJ, Tijms BM, van Berckel BNM, Le Bastard N, van der Flier WM, Scheltens P, et al. Comparing CSF amyloid-beta biomarker ratios for two automated immunoassays, Elecsys and Lumipulse, with amyloid PET status. Alzheimers Dement (Amst). 2021;13:e12182.

[19] Chong JR, Ashton NJ, Karikari TK, Tanaka T, Saridin FN, Reilhac A, et al. Plasma P-tau181 to Abeta42 ratio is associated with brain amyloid burden and hippocampal atrophy in an Asian cohort of Alzheimer’s disease patients with concomitant cerebrovascular disease. Alzheimers Dement. 2021;17:1649–62.

[20] Fowler C, Rainey-Smith SR, Bird S, Bomke J, Bourgeat P, Brown BM, et al. Fifteen Years of the Australian Imaging, Biomarkers and Lifestyle (AIBL) Study: Progress and Observations from 2,359 Older Adults Spanning the Spectrum from Cognitive Normality to Alzheimer’s Disease. J Alzheimers Dis Rep. 2021;5:443–68.

[21] Bransby L, Lim YY, Ames D, Fowler C, Roberston J, Harrington K, et al. Sensitivity of a Preclinical Alzheimer’s Cognitive Composite (PACC) to amyloid beta load in preclinical Alzheimer’s disease. J Clin Exp Neuropsychol. 2019;41:591–600.

[22] Burnham SC, Bourgeat P, Dore V, Savage G, Brown B, Laws S, et al. Clinical and cognitive trajectories in cognitively healthy elderly individuals with suspected non-Alzheimer’s disease pathophysiology (SNAP) or Alzheimer’s disease pathology: a longitudinal study. Lancet Neurol. 2016;15:1044–53.

[23] Bourgeat P, Doré V, Fripp J, Ames D, Masters CL, Salvado O, et al. Implementing the centiloid transformation for 11c-PiB and β-amyloid 18f-PET tracers using CapAIBL. NeuroImage. 2018;183:387–93.

[24] Klunk WE, Koeppe RA, Price JC, Benzinger TL, Devous MD, Sr., Jagust WJ, et al. The Centiloid Project: standardizing quantitative amyloid plaque estimation by PET. Alzheimers Dement. 2015;11:1-15.e1-4.

[25] Rowe CC, Jones G, Dore V, Pejoska S, Margison L, Mulligan RS, et al. Standardized expression of 18F-NAV4694 and 11C-PiB β-amyloid PET results with the Centiloid Scale. Journal of Nuclear Medicine. 2016;57:1233–7.

[26] Krishnadas N, Villemagne VL, Dore V, Rowe CC. Advances in Brain Amyloid Imaging. Semin Nucl Med. 2021;51:241–52.

[27] Amadoru S, Dore V, McLean CA, Hinton F, Shepherd CE, Halliday GM, et al. Comparison of amyloid PET measured in Centiloid units with neuropathological findings in Alzheimer’s disease. Alzheimers Res Ther. 2020;12:22.

[28] La Joie R, Ayakta N, Seeley WW, Borys E, Boxer AL, DeCarli C, et al. Multisite study of the relationships between antemortem [(11)C]PIB-PET Centiloid values and postmortem measures of Alzheimer’s disease neuropathology. Alzheimers Dement. 2019;15:205–16.

[29] Thijssen EH, Verberk IMW, Vanbrabant J, Koelewijn A, Heijst H, Scheltens P, et al. Highly specific and ultrasensitive plasma test detects Abeta(1-42) and Abeta(1-40) in Alzheimer’s disease. Sci Rep. 2021;11:9736.

[30] Bayoumy S, Verberk IMW, den Dulk B, Hussainali Z, Zwan M, van der Flier WM, et al. Clinical and analytical comparison of six Simoa assays for plasma P-tau isoforms P-tau181, P- tau217, and P-tau231. Alzheimers Res Ther. 2021;13:198.

[31] Chong JR, Ashton NJ, Karikari TK, Tanaka T, Scholl M, Zetterberg H, et al. Blood-based high sensitivity measurements of beta-amyloid and phosphorylated tau as biomarkers of Alzheimer’s disease: a focused review on recent advances. J Neurol Neurosurg Psychiatry. 2021;92:1231–41.

[32] Verberk IMW, Slot RE, Verfaillie SCJ, Heijst H, Prins ND, van Berckel BNM, et al. Plasma Amyloid as Prescreener for the Earliest Alzheimer Pathological Changes. Ann Neurol. 2018;84:648–58.

[33] De Meyer S, Schaeverbeke JM, Verberk IMW, Gille B, De Schaepdryver M, Luckett ES, et al. Comparison of ELISA- and SIMOA-based quantification of plasma Abeta ratios for early detection of cerebral amyloidosis. Alzheimers Res Ther. 2020;12:162.

[34] Schindler SE, Bollinger JG, Ovod V, Mawuenyega KG, Li Y, Gordon BA, et al. High-precision plasma beta-amyloid 42/40 predicts current and future brain amyloidosis. Neurology. 2019;93:e1647–e59.

[35] Sjogren M, Vanderstichele H, Agren H, Zachrisson O, Edsbagge M, Wikkelso C, et al. Tau and Abeta42 in cerebrospinal fluid from healthy adults 21-93 years of age: establishment of reference values. Clin Chem. 2001;47:1776–81.

[36] Wu X, Xiao Z, Yi J, Ding S, Gu H, Wu W, et al. Development of a Plasma Biomarker Diagnostic Model Incorporating Ultrasensitive Digital Immunoassay as a Screening Strategy for Alzheimer Disease in a Chinese Population. Clin Chem. 2021;67:1628–39.

[37] Harari O, Cruchaga C, Kauwe JS, Ainscough BJ, Bales K, Pickering EH, et al. Phosphorylated tau-Abeta42 ratio as a continuous trait for biomarker discovery for early-stage Alzheimer’s disease in multiplex immunoassay panels of cerebrospinal fluid. Biol Psychiatry. 2014;75:723–31.

