## Supplementary Information for "Plasma p-tau181/Aβ_1-42_ ratio predicts Aβ-PET status and correlates with CSF-p-tau181/Aβ_1-42_ and future cognitive decline"

**METHODOLOGY**

***AIBL longitudinal cognitive information***

Longitudinal cognitive information was aligned such that the baseline cognitive measure was the baseline plasma collection. Sample sizes for the number of samples used in the assessment of cognitive decline both in the whole cohort (WC) and in the Aβ-PET groups (using the CL threshold at 20CL) are shown in Supplementary table 1.

***Sample analysis***

A total of 387 EDTA plasma samples were analyzed in duplicate in 15 consecutive test runs over 13 days on two Simoa HD-X devices. Two re-runs were performed for re-analysis of samples with missing values or too high variation in the duplicate results (%CV>20 for Amyloid, %CV>25 for p-tau). Samples with a concentration below the LLOQ were not considered for retesting. The samples were stored at -80°C at the testing site (ADx NeuroSciences) and thawed on the bench prior to use in the assay runs (single freeze-thaw cycle). The samples in the re-runs had an additional freeze-thaw cycle. After thawing, samples were centrifuged (10,000xg for 8 minutes) and pre-diluted manually in the 96-well polypropylene measurement plate (supplementary table 2 for more details) before analysis in the Simoa HD-X instrument. Furthermore a QC panel was composed separately for the plasma Amyloid and for the pTau assays. After sample screening (individual healthy donor samples), 2 samples were selected for each assay and further aliquoted into polypropylene test tubes (Sarstedt Cat n° 72730105; 120 µL/vial), frozen at -20°C and thawed upon testing in each assay run to monitor run-to-run variability.

***QC panel testing and run to run variability***

The data of the QC panel are reported in Supplementary data Table 11. While the panel for p-tau assays suffered from too low signals in this study and was not further explored (data not shown) the 2 QC samples for the Amyloid assays showed concentrations within the measuring range of the methods. The $A\beta_{1\text{-}40}$ and respectively $A\beta_{1\text{-}42}$ median concentration (not corrected for the dilution factor) for both controls was 5.51 /7.43 pg/mL (QC1/QC2) and 3.84 /5.57 pg/mL (QC1/QC2). The variation over the runs was respectively 5.8/5.7 %CV for $A\beta_{1\text{-}40}$ and 7.1/13.5 %CV for $A\beta_{1\text{-}42}$.

In addition to the control panel the variation on the calibrator points was investigated (Supplementary data Table 12). A median inter-run variation of respectively 8.4, 9.0, 9.2 and 14.1 %CV was reported for the $A\beta_{1\text{-}40}$, $A\beta_{1\text{-}42}$, p-tau181 and p-tau231 over the 7 non-zero calibrator points. This low variation confirms the findings of the QC panel. Given the low variation for both QC panel and calibrator points (below %CV 15) no correction was required for variation or trending nor for Amyloid nor for p-tau assays. In line with the expectations we found no samples below LLOQ in both plasma Amyloid assays compared to the p-tau assays. There were 9 samples below the LLOQ for p-tau181 assay and 6 samples for the p-tau231 assay. A similar trend was observed for the repeatability. There was a single sample above %CV 20 for both Amyloid tests and respectively 21 and 32 samples for p-tau181 and p-tau231 (Supplementary Table 13).

***Accuracy and intra-run precision***

The accuracy of the assay methods was assessed by back calculation of the calibrator point concentrations (supplemental data table 14) and expressed as a percentage of the theoretical concentration. For all 4 assays the accuracy was between 80-120% with the exception of the lowest calibrator points of the p-tau181 and p-tau231 assay that were respectively 130.9 % (0.78 pg/mL) and 122.0% (0.39 pg/mL). The assays also demonstrated an acceptable intra-assay variation that is illustrated by the median variation on the duplicate measurements (supplementary table 15). The percentage variation was respectively 2.20, 2.14, 6.13 and 7.00 %CV for the $A\beta_{1-40}$, $A\beta_{1-42}$, p-tau181 and p-tau231 Simoa assay.

***Statistical methodology***

Demographic characteristics were assessed between Aβ-PET groups using a combination of unpaired t-tests for parametric quantitative variables (age, the AIBL PACC, CL and CDR SOB), Kruskall-Wallis test for MMSE and Chi-square test for comparison of categorical variables such as *APOE* ε4 allele status, clinical classification and tracer.

Comparison of the marginal means between Aβ-PET groups was performed using Generalised Linear modelling (GLM, binomial link), with models adjusted for age, gender, *APOE* ε4 allele status and tracer. Aβ-PET group prediction statistics were performed using Receiver Operating Characteristic (ROC) analyses, with Area under the curve (AUC), sensitivity, specificity, accuracy, negative predictive values (NPV) and positive predictive values (PPV) calculated for both individual plasma biomarkers, and model fits given the combination of plasma biomarker with confounders (age, gender, *APOE* ε4 allele status and tracer). Thresholds were derived using Youden's Index. ROC models consisting of the plasma biomarker and study confounders were compared using DeLong's method with a model containing the confounders only (base model) to assess whether the addition of each plasma marker was significantly better than the base model alone. CSF $A\beta_{1-42}$ group prediction statistics were performed in the same fashion as the Aβ-PET group prediction statistics, with $A\beta_{1-42}$ groups derived using the threshold from Doecke *et al.* (2020) at 1054 pg/µL.

Multivariate modelling was performed using the stepAIC function in R including all plasma biomarkers and confounders. The process removes features in the model in a stepwise fashion until the most parsimonious model is derived. Final model was compared then to the best performing model containing only one biomarker.

Correlation between plasma and CSF markers and plasma and CL values was performed using Spearman's Rho ($\rho$).

Change in cognition using only the best performing plasma marker along with confounders age, gender, *APOE* $\varepsilon$4 allele status and tracer using linear mixed effects models (LME) using both a random intercept and a random slope. Two versions were considered, one assessing the interaction between time and the quantitative plasma marker, and two assessing the interaction between time the categorical version of the plasma marker as derived by Youden's Index. To assess the performance of comparable CSF biomarkers to predict cognitive decline, the same statistical design using LME models was performed.

To assess the associations across different cognitive groups, mean plasma biomarker comparisons and ROC analyses were performed in both the total cohort and amongst cognitively normal (CN) and cognitively impaired (CI) groups.

All statistical models were computed using the complete sample at both visits 1 & 2 including the imputed Aβ-PET status. Further ROC models were performed at the three designated CL thresholds using the non-imputed data, with results shown in supplementary information.

P-values were compared against a Bonferroni adjusted alpha ($\alpha$ = 0.05/6 = 0.008). All statistical analyses were performed using the R statistical environment.

Supplementary Table 1: Sample size for longitudinal assessment of cognition.

| **Collection** | **Plasma N WC** | **Plasma CN** | **Plasma CI** | **CSF N WC** | **CSF CN** | **CSF CI** |
|---|---|---|---|---|---|---|
| Baseline | 233 | 168 | 65 | 155 | 110 | 45 |
| 18 months | 217 | 153 | 64 | 148 | 99 | 49 |
| 36 months | 162 | 120 | 42 | 114 | 82 | 32 |
| 54 months | 124 | 105 | 19 | 86 | 71 | 15 |
| 72 months | 101 | 89 | 12 | 73 | 64 | 9 |
| 90 months | 66 | 60 | 6 | 45 | 41 | 4 |
| 108 months | 36 | 35 | 1 | 24 | 24 | 0 |
| 124 months | 13 | 13 | 0 | 8 | 8 | 0 |
| 144 months | 3 | 3 | 0 | 1 | 1 | 0 |

Supplementary Table 2: Detailed test specifications of the A$\beta_{1-40}$, A$\beta_{1-42}$, p-tau181 and p-tau231 assay.

<table>
<tr><td colspan="3"></td><td colspan="2">Plasma Amyloid assays</td></tr>
<tr><td colspan="3">ANALYTE</td><td>A$\beta_{1-40}$</td><td>A$\beta_{1-42}$</td></tr>
<tr><td colspan="3">REGULATORY STATUS</td><td colspan="2">Prototype</td></tr>
<tr><td colspan="3">MATRIX</td><td colspan="2">EDTA plasma</td></tr>
<tr><td colspan="3">TECHNOLOGY</td><td colspan="2">Quanterix Simoa platform HDx / Single plex assays</td></tr>
<tr><td rowspan="8">BIOMATERIALS</td><td rowspan="3">Capture mAb</td><td>Name</td><td>ADx103</td><td>ADx102</td></tr>
<tr><td>Species/Isotype</td><td>Mouse IgG1</td><td>Mouse IgG2a</td></tr>
<tr><td>Epitope</td><td>Aβ x-40</td><td>Aβ x-42</td></tr>
<tr><td rowspan="3">Detector mAb</td><td>Name</td><td colspan="2">ADx101</td></tr>
<tr><td>Species/Isotype</td><td colspan="2">IgG2b</td></tr>
<tr><td>Epitope</td><td colspan="2">Aβ 1-x</td></tr>
<tr><td rowspan="2">Calibrator</td><td>Type</td><td colspan="2">Recombinant protein (E.coli)</td></tr>
<tr><td>Source</td><td>rPeptide Cat n°A-1153-1</td><td>rPeptide Cat n°A-1163-1</td></tr>
<tr><td rowspan="17">ASSAY</td><td rowspan="8">Sample incubation</td><td>Simultaneous/<br>sequential with detector mAb</td><td colspan="2">Simultaneous</td></tr>
<tr><td>Volume diluted sample/cuvet (µL)</td><td colspan="2">100</td></tr>
<tr><td>Volume Detector/cuvet (µL)</td><td colspan="2">20</td></tr>
<tr><td>Volume Beads/cuvet (µL)</td><td colspan="2">25</td></tr>
<tr><td>Total volume/cuvet</td><td colspan="2">145</td></tr>
<tr><td>Sample pre-dilution</td><td>20x</td><td>4x</td></tr>
<tr><td>% Plasma/cuvet</td><td>3.4</td><td>17.2</td></tr>
<tr><td>Recipient for pre-dilution</td><td colspan="2">PP (96 well conical microtiter-plate)<br>(Thermo Scientific Cat#249944)</td></tr>
<tr><td rowspan="4">Boundary Conditions</td><td rowspan="2">Sample/detector incubation</td><td colspan="2">80 cadences (60 minutes)</td></tr>
<tr><td colspan="2">Room temperature</td></tr>
<tr><td rowspan="2">SβG conjugate incubation</td><td colspan="2">7 cadences (5.5 minutes)</td></tr>
<tr><td colspan="2">Room temperature</td></tr>
<tr><td rowspan="5">Calibrators</td><td>Format</td><td colspan="2">Ready-to-use</td></tr>
<tr><td>Design</td><td colspan="2">Liquid; stored at -20°C and thawed prior to use</td></tr>
<tr><td>Number</td><td colspan="2">7 non-zero + BLANK</td></tr>
<tr><td>Concentration range (pg/mL)</td><td>1-20</td><td>0.98-18.4</td></tr>
<tr><td>Curve-fit algorithm</td><td colspan="2">4PL non weighing</td></tr>
</table>

| | | | Plasma p-tau assays | |
|---|---|---|---|---|
| **ANALYTE** | | | p-tau181 | p-tau231 |
| REGULATORY STATUS | | | Prototype | |
| MATRIX | | | EDTA plasma | |
| TECHNOLOGY | | | Quanterix Simoa platform HDx / Single plex assays | |
| BIOMATERIALS | Capture mAb | Name | ADx252 | ADx253 |
| | | Species/Isotype | Rabbit IgG | Mouse IgG2b |
| | | Epitope | pT181 | pT231 |
| | Detector mAb | Name | ADx204 | |
| | | Species/Isotype | Mouse IgG1 | |
| | | Epitope | N-terminal | |
| | Calibrator | Type | Synthetic peptide covering epitopes of ADx204, ADx252 and ADx253 (6116.78 Da) | |
| | | Source | Proteogenix | |
| ASSAY | Sample incubation | Simultaneous/<br>sequential with detector mAb | Simultaneous | |
| | | Volume diluted sample/cuvet (μL) | 100 | |
| | | Volume Detector/cuvet (μL) | 20 | |
| | | Volume Beads/cuvet (μL) | 25 | |
| | | Total volume/cuvet (μL) | 145 | |
| | | Sample pre-dilution | 5x | |
| | | % Plasma/cuvet | 13.8 | |
| | | Recipient for pre-dilution | PP (96 well conical microtiter-plate)<br>(Thermo Scientific Cat#249944) | |
| | Boundary Conditions | Sample/detector incubation | 80 cadences (60 minutes) | |
| | | | Room temperature | |
| | | SβG conjugate incubation | 14 cadences (10 minutes) | 7 cadenses (5.5 minutes) |
| | | | Room temperature | |
| | Calibrators | Format | Ready-to-use | |
| | | Design | Liquid; stored at -20°C and thawed prior to use | |
| | | Number | 7 non-zero + BLANK | |
| | | Concentration range (pg/mL) | 0.78-50 | 0.39-50 |
| | | Curve-fit algorithm | $1/y^2$ weighted 5PL including blank AEB (set at 0.1 pg/mL) | |

Supplementary Table 3: Study demographic characteristics at Assessment 2

| | **Total Sample** | **A$\beta^-$** | **A$\beta^+$** | **p-value** |
|---|---|---|---|---|
| N (%) | 100 | 62 (62%) | 38 (38%) | |
| Gender Male, N (%) | 51 (51%) | 30 (48%) | 21 (55%) | 0.5 |
| Mean Age, years (SD) | 75.8 (6.3) | 75.4 (6.8) | 76.4 (5.4) | 0.4 |
| *APOE* ε4 Carriage, N (%) | 25 (25%) | 7 (11%) | 18 (47%) | <0.0001 |
| Tracer Florbetapir N % | 21 (21%) | 16 (21%) | 5 (21%) | |
| Tracer Flutemetamol N % | 31 (31%) | 18 (31%) | 13 (31%) | |
| Tracer NAV/PiB N % | 48 (48%) | 28 (48%) | 20 (48%) | 0.36 |
| Mean CL (SD) | 29.3 (42.8) | 1.8 (8.2) | 76.3 (36.5) | <0.0001 |
| Diagnosis CN N % | 78 (78%) | 56 (56%) | 22 (22%) | |
| Diagnosis MCI N % | 14 (14%) | 5 (5%) | 9 (9%) | |
| Diagnosis AD N % | 8 (8%) | 1 (1%) | 7 (7%) | 0.00041 |
| Median MMSE, (MAD) | 29 (1.5) | 29 (1.5) | 28 (3) | 0.055 |
| Median CDR-SOB, (MAD) | 0 (0) | 0 (0) | 0.5 (0.7) | 0.00018 |
| Mean AIBL PACC (SD) | 100 | 62 (62%) | 38 (38%) | |

Characteristics measured at Assessment 2, and compared between Aβ-PET groups using a CL threshold at 20CL.

Supplementary Table 4: Mean biomarker comparisons, Assessments 1 & 2, using CL threshold 20

| Biomarker | Visit | N Aβ- | N Aβ+ | Mean Aβ- | Mean Aβ+ | Median Aβ- | Median Aβ+ | p-value[1] | p-value[2] |
|---|---|---|---|---|---|---|---|---|---|
| $A\beta_{1-40}$ | 1 | 142 | 91 | 150 (20) pg/mL | 151 (20.8) pg/mL | 150 (21.3) pg/mL | 148 (18.5) pg/mL | 8.15E-01 | 5.97E-01 |
| $A\beta_{1-42}$ | 1 | 142 | 91 | 20.1 (4.09) pg/mL | 17.2 (3.33) pg/mL | 19.8 (4.03) pg/mL | 17.6 (3.45) pg/mL | 2.14E-08 | 2.06E-05 |
| $A\beta_{1-42/1-40}$ | 1 | 142 | 91 | 0.135 (0.0231) | 0.115 (0.0212) | 0.136 (0.0242) | 0.117 (0.0169) | 7.95E-11 | 8.26E-06 |
| p-tau181 | 1 | 142 | 91 | 17 (11.7) pg/mL | 37.2 (16.6) pg/mL | 14.6 (6.27) pg/mL | 36 (14.1) pg/mL | 2.24E-23 | 7.55E-12 |
| p-tau231 | 1 | 142 | 91 | 8.09 (4.16) pg/mL | 10.3 (4.45) pg/mL | 7.48 (3.45) pg/mL | 9.49 (4.05) pg/mL | 3.81E-05 | 1.26E-02 |
| p-tau181/$A\beta_{1-42}$ | 1 | 142 | 91 | 0.875 (0.64) | 2.24 (1.05) | 0.739 (0.375) | 2.04 (0.994) | 2.01E-27 | 7.86E-13 |
| p-tau231/$A\beta_{1-42}$ | 1 | 142 | 91 | 0.418 (0.234) | 0.639 (0.389) | 0.39 (0.19) | 0.536 (0.231) | 1.02E-08 | 1.73E-04 |
| $A\beta_{1-40}$ | 2 | 62 | 38 | 153 (15.7) pg/mL | 147 (16.2) pg/mL | 153 (16.1) pg/mL | 144 (13.2) pg/mL | 8.69E-02 | 6.84E-03 |
| $A\beta_{1-42}$ | 2 | 62 | 38 | 20.7 (3.83) pg/mL | 16.8 (3.62) pg/mL | 20.8 (2.95) pg/mL | 17.8 (2.62) pg/mL | 1.55E-06 | 2.24E-04 |
| $A\beta_{1-42/1-40}$ | 2 | 62 | 38 | 0.136 (0.0214) | 0.112 (0.0228) | 0.14 (0.0218) | 0.12 (0.0141) | 2.36E-06 | 1.15E-03 |
| p-tau181 | 2 | 62 | 38 | 15.5 (9.35) pg/mL | 39.8 (18.7) pg/mL | 14 (6.63) pg/mL | 38.9 (22.1) pg/mL | 2.59E-13 | 5.37E-06 |
| p-tau231 | 2 | 62 | 38 | 8.54 (4.75) pg/mL | 10.8 (4.41) pg/mL | 7.36 (3.53) pg/mL | 9.81 (3.57) pg/mL | 3.31E-03 | 1.14E-01 |
| p-tau181/$A\beta_{1-42}$ | 2 | 62 | 38 | 0.758 (0.459) | 2.56 (1.57) | 0.694 (0.294) | 2.29 (1.3) | 1.60E-15 | 7.47E-06 |
| p-tau231/$A\beta_{1-42}$ | 2 | 62 | 38 | 0.425 (0.244) | 0.692 (0.373) | 0.367 (0.21) | 0.604 (0.243) | 1.18E-05 | 4.42E-03 |

***Biomarker mean comparisons between Aβ-PET groups***

Using CL thresholds at 15, 20 and 25 to define Aβ-PET groups did not change which plasma markers were significantly altered in the Aβ-PET$^+$ group as compared with Aβ-PET$^-$ group. Supplementary Table 4 shows mean values per Aβ-PET group (CL threshold at 20 CL), with p-values both unadjusted and adjusted (for confounders). Figures 1A and 1B show the each of the markers plotted against Aβ-PET groups using the CL threshold at 20CL. All markers except for $A\beta_{1-40}$ and p-tau231 were significantly altered in the Aβ-PET$^+$ group ($P<0.008$). Ratio markers $A\beta_{1-42/1-40}$ and p-tau181/$A\beta_{1-42}$ showed slightly stronger differences between Aβ-PET group in comparison with $A\beta_{1-42}$ and p-tau181 alone. The p-tau231/$A\beta_{1-42}$ was significantly higher in the Aβ-PET$^{+}$ group ($P=0.0003$), however the difference was not as strong as the same comparison for p-tau181/$A\beta_{1-42}$ ($P<0.00001$).

Supplementary Table 5: Unadjusted AUC values from individual biomarkers across two Assessments and CL thresholds 15, 20 & 25

| Biomarker | Threshold | Complete sample<br>Assessment 1<br>AUC (95%CI) | Assessment 2<br>AUC (95%CI) | CN<br>Assessment 1<br>AUC (95%CI) | Assessment 2<br>AUC (95%CI) | CI<br>Assessment 1<br>AUC (95%CI) | Assessment 2<br>AUC (95%CI) |
|---|---|---|---|---|---|---|---|
| N (Aβ-/Aβ+) | | 128/105 | 56/44 | 113/55 | 51/27 | 15/50 | 5/17 |
| $A\beta_{1\text{-}42}$ | @ CL15 | 0.681 (0.61 – 0.75) | 0.755 (0.66 – 0.85) | 0.667 (0.58 – 0.75) | 0.741 (0.63 – 0.85) | 0.738 (0.59 – 0.89) | 0.871 (0.61 – 1) |
| $A\beta_{1\text{-}42/1\text{-}40}$ | | 0.721 (0.66 - 0.79) | 0.745 (0.65 - 0.84) | 0.679 (0.59 - 0.76) | 0.741 (0.63 - 0.85) | 0.872 (0.75 - 0.99) | 0.847 (0.68 - 1) |
| p-tau181 | | 0.841 (0.79 - 0.9) | 0.851 (0.77 - 0.93) | 0.782 (0.7 - 0.86) | 0.802 (0.7 - 0.91) | 0.848 (0.71 - 0.99) | 0.918 (0.79 - 1) |
| p-tau231 | | 0.665 (0.6 - 0.74) | 0.632 (0.52 - 0.74) | 0.666 (0.58 - 0.75) | 0.602 (0.47 - 0.73) | 0.607 (0.42 - 0.8) | 0.459 (0.12 - 0.8) |
| **p-tau181/$A\beta_{1\text{-}42}$** | | **0.859 (0.81 - 0.91)** | **0.883 (0.81 - 0.95)** | **0.798 (0.72 - 0.88)** | **0.843 (0.74 - 0.94)** | **0.892 (0.78 - 1)** | **0.965 (0.89 - 1)** |
| p-tau231/$A\beta_{1\text{-}42}$ | | 0.709 (0.64 - 0.78) | 0.698 (0.6 - 0.8) | 0.702 (0.62 - 0.79) | 0.679 (0.56 - 0.8) | 0.657 (0.48 - 0.83) | 0.718 (0.42 - 1) |
| N (Aβ-/Aβ+) | | 142/91 | 62/38 | 124/44 | 56/22 | 18/47 | 6/16 |
| $A\beta_{1\text{-}42}$ | @ CL20 | 0.705 (0.64 - 0.77) | 0.793 (0.71 - 0.88) | 0.705 (0.62 - 0.79) | 0.781 (0.67 - 0.89) | 0.741 (0.6 - 0.88) | 0.896 (0.69 - 1) |
| $A\beta_{1\text{-}42/1\text{-}40}$ | | 0.731 (0.69 - 0.81) | 0.787 (0.7 - 0.87) | 0.728 (0.64 - 0.81) | 0.786 (0.68 - 0.89) | 0.852 (0.74 - 0.96) | 0.906 (0.78 - 1) |
| p-tau181 | | 0.862 (0.81 - 0.92) | 0.904 (0.84 - 0.97) | 0.808 (0.72 - 0.89) | 0.862 (0.77 - 0.96) | 0.868 (0.75 - 0.98) | 0.958 (0.87 - 1) |
| p-tau231 | | 0.66 (0.59 - 0.73) | 0.673 (0.57 - 0.78) | 0.646 (0.55 - 0.74) | 0.667 (0.54 - 0.79) | 0.636 (0.47 - 0.8) | 0.458 (0.17 - 0.75) |
| **p-tau181/$A\beta_{1\text{-}42}$** | | **0.883 (0.83 - 0.93)** | **0.942 (0.89 - 1)** | **0.832 (0.75 - 0.91)** | **0.912 (0.83 - 1)** | **0.908 (0.82 - 1)** | **1 (1 - 1)** |
| p-tau231/$A\beta_{1\text{-}42}$ | | 0.713 (0.65 - 0.78) | 0.749 (0.65 - 0.85) | 0.692 (0.6 - 0.78) | 0.751 (0.64 - 0.87) | 0.696 (0.54 - 0.85) | 0.75 (0.5 - 1) |
| N (Aβ-/Aβ+) | | 146/87 | 66/34 | 128/40 | 60/18 | 18/47 | 6/16 |
| $A\beta_{1\text{-}42}$ | @ CL25 | 0.71 (0.64 - 0.78) | 0.769 (0.68 - 0.86) | 0.719 (0.64 - 0.8) | 0.757 (0.64 - 0.88) | 0.741 (0.6 - 0.88) | 0.896 (0.69 - 1) |
| $A\beta_{1\text{-}42/1\text{-}40}$ | | 0.762 (0.7 - 0.82) | 0.785 (0.7 - 0.87) | 0.753 (0.68 - 0.83) | 0.795 (0.69 - 0.9) | 0.852 (0.74 - 0.96) | 0.906 (0.78 - 1) |
| p-tau181 | | 0.886 (0.84 - 0.94) | 0.955 (0.92 - 0.99) | 0.855 (0.78 - 0.93) | 0.946 (0.9 - 0.99) | 0.868 (0.75 - 0.98) | 0.958 (0.87 - 1) |
| p-tau231 | | 0.654 (0.58 - 0.73) | 0.722 (0.62 - 0.82) | 0.639 (0.54 - 0.74) | 0.746 (0.63 - 0.86) | 0.636 (0.47 - 0.8) | 0.458 (0.17 - 0.75) |
| **p-tau181/$A\beta_{1\text{-}42}$** | | **0.905 (0.86 - 0.95)** | **0.97 (0.94 - 1)** | **0.873 (0.8 - 0.94)** | **0.958 (0.92 - 1)** | **0.908 (0.82 - 1)** | **1 (1 - 1)** |
| p-tau231/$A\beta_{1\text{-}42}$ | | 0.712 (0.65 - 0.78) | 0.776 (0.68 - 0.87) | 0.693 (0.6 - 0.79) | 0.801 (0.7 - 0.91) | 0.696 (0.54 - 0.85) | 0.75 (0.5 - 1) |

***p-tau181/A$\beta_{1\text{-}42}$ ratio performs better than the p-tau181 alone***

Comparing the performance of the individual p-tau181 marker to that of the p-tau181/A$\beta_{1\text{-}42}$ ratio to predict Aβ-PET status (in biomarker models only [without confounders] at Assessment 1) found that at the lower CL threshold (15CL) the AUC for the ratio was not significantly greater than that for the individual p-tau181 (P=0.0849). At both higher thresholds (20CL & 25CL) however, the ratio outperformed the p-tau181 marker alone (@20CL P=0.0156 & @ CL25 P=0.0183, supplementary table 5). At Assessment 2, comparative P-values for the ROC models at CL thresholds 15, 20 and 25 were similar, except for the higher CL threshold (@15CL P=0.0546, @20CL P=0.0185, @25CL P=0.0934, Supplementary Table 5).

Supplementary Table 6: AUC values from biomarkers across two Assessments and CL thresholds 15, 20 & 25 using non-imputed Aβ-PET

| Biomarker | Threshold | Complete sample<br>Assessment 1<br>AUC (95%CI) | <br>Assessment 2<br>AUC (95%CI) | CN<br>Assessment 1<br>AUC (95%CI) | <br>Assessment 2<br>AUC (95%CI) | CI<br>Assessment 1<br>AUC (95%CI) |
|---|---|---|---|---|---|---|
| N (Aβ-/Aβ+) | | 88/81 | 40/25 | 75/40 | 37/13 | 13/41 |
| $A\beta_{1-42}$ | @ CL15 | 0.719 (0.64 - 0.8) | 0.742 (0.62 - 0.86) | 0.71 (0.61 - 0.81) | 0.742 (0.58 - 0.9) | 0.744 (0.57 - 0.92) |
| $A\beta_{1-42/1-40}$ | | 0.742 (0.67 - 0.82) | 0.739 (0.62 - 0.86) | 0.711 (0.62 - 0.81) | 0.723 (0.57 - 0.88) | 0.857 (0.72 - 0.99) |
| p-tau181 | | 0.845 (0.78 - 0.91) | 0.884 (0.8 - 0.97) | 0.793 (0.7 - 0.89) | 0.796 (0.67 - 0.93) | 0.878 (0.75 - 1) |
| p-tau231 | | 0.638 (0.55 - 0.72) | 0.662 (0.53 - 0.8) | 0.649 (0.54 - 0.75) | 0.57 (0.39 - 0.75) | 0.576 (0.36 - 0.79) |
| p-tau181/$A\beta_{1-42}$ | | 0.869 (0.81 - 0.93) | 0.922 (0.85 - 0.99) | 0.817 (0.73 - 0.91) | 0.867 (0.75 - 0.98) | 0.923 (0.84 - 1) |
| p-tau231/$A\beta_{1-42}$ | | 0.695 (0.62 - 0.77) | 0.732 (0.61 - 0.86) | 0.71 (0.61 - 0.81) | 0.672 (0.49 - 0.85) | 0.364 (0.16 - 0.56) |
| N (Aβ-/Aβ+) | | 99/70 | 41/24 | 83/32 | 38/12 | 16/38 |
| $A\beta_{1-42}$ | @ CL20 | 0.732 (0.66 - 0.81) | 0.77 (0.66 - 0.88) | 0.727 (0.62 - 0.83) | 0.789 (0.65 - 0.93) | 0.749 (0.59 - 0.9) |
| $A\beta_{1-42/1-40}$ | | 0.77 (0.7 - 0.84) | 0.766 (0.65 - 0.88) | 0.762 (0.67 - 0.85) | 0.768 (0.63 - 0.91) | 0.837 (0.71 - 0.96) |
| p-tau181 | | 0.872 (0.81 - 0.93) | 0.902 (0.83 - 0.98) | 0.837 (0.74 - 0.93) | 0.827 (0.71 - 0.95) | 0.891 (0.79 - 1) |
| p-tau231 | | 0.64 (0.56 - 0.72) | 0.688 (0.56 - 0.82) | 0.643 (0.53 - 0.76) | 0.61 (0.43 - 0.79) | 0.613 (0.43 - 0.8) |
| p-tau181/$A\beta_{1-42}$ | | 0.895 (0.84 - 0.95) | 0.947 (0.9 - 1) | 0.86 (0.77 - 0.95) | 0.914 (0.84 - 0.99) | 0.933 (0.86 - 1) |
| p-tau231/$A\beta_{1-42}$ | | 0.699 (0.62 - 0.78) | 0.76 (0.64 - 0.88) | 0.697 (0.59 - 0.8) | 0.719 (0.55 - 0.89) | 0.679 (0.51 - 0.85) |
| N (Aβ-/Aβ+) | | 101/68 | 44/21 | 85/30 | 41/9 | 16/38 |
| $A\beta_{1-42}$ | @ CL25 | 0.733 (0.66 - 0.81) | 0.719 (0.6 - 0.84) | 0.736 (0.64 - 0.83) | 0.729 (0.56 - 0.9) | 0.749 (0.59 - 0.9) |
| $A\beta_{1-42/1-40}$ | | 0.772 (0.7 - 0.84) | 0.742 (0.62 - 0.87) | 0.771 (0.69 - 0.86) | 0.74 (0.57 - 0.91) | 0.837 (0.71 - 0.96) |
| p-tau181 | | 0.886 (0.83 - 0.95) | 0.947 (0.89 - 1) | 0.866 (0.78 - 0.96) | 0.911 (0.83 - 0.99) | 0.891 (0.79 - 1) |
| p-tau231 | | 0.626 (0.54 - 0.71) | 0.742 (0.62 - 0.86) | 0.618 (0.5 - 0.74) | 0.699 (0.52 - 0.88) | 0.613 (0.43 - 0.8) |
| p-tau181/$A\beta_{1-42}$ | | 0.906 (0.85 - 0.96) | 0.957 (0.91 - 1) | 0.882 (0.8 - 0.96) | 0.927 (0.85 - 1) | 0.933 (0.86 - 1) |
| p-tau231/$A\beta_{1-42}$ | | 0.69 (0.61 - 0.77) | 0.78 (0.66 - 0.9) | 0.684 (0.58 - 0.79) | 0.756 (0.57 - 0.94) | 0.679 (0.51 - 0.85) |

ROC statistics not shown for Assessment 2 in the CI group as there were only 3 Aβ-PET- and 12 Aβ-PET+ participants.

Supplementary Table 7: AUC values from ROC models adjusted for age, gender and *APOE* ε4 allele status

| Biomarker | Threshold | Complete sample<br>Assessment 1<br>AUC (95%CI) | Assessment 2<br>AUC (95%CI) | CN<br>Assessment 1<br>AUC (95%CI) | Assessment 2<br>AUC (95%CI) | CI<br>Assessment 1<br>AUC (95%CI) |
|---|---|---|---|---|---|---|
| N ($A\beta^-/A\beta^+$) | @ CL15 | 128/105 | 56/44 | 113/55 | 51/27 | 15/50 |
| Base model | | 0.739 (0.68 - 0.8) | 0.738 (0.64 - 0.84) | 0.718 (0.64 - 0.8) | 0.774 (0.67 - 0.88) | 0.777 (0.65 - 0.91) |
| $A\beta_{1-42}$ | | 0.771 (0.71 - 0.83) | 0.784 (0.7 - 0.87) | 0.740 (0.66 - 0.82) | 0.807 (0.71 - 0.9) | 0.888 (0.79 - 0.98) |
| $A\beta_{1-42/1-40}$ | | 0.779 (0.72 - 0.84)* | 0.772 (0.68 - 0.86) | 0.740 (0.66 - 0.82) | 0.79 (0.69 - 0.89) | 0.941 (0.87 - 1)* |
| p-tau181 | | 0.865 (0.82 - 0.91)*** | 0.872 (0.80 - 0.94)*** | 0.819 (0.75 - 0.89)* | 0.844 (0.75 - 0.93) | 0.908 (0.83 - 0.99) |
| p-tau231 | | 0.766 (0.7 - 0.83) | 0.758 (0.66 - 0.85) | 0.753 (0.67 - 0.83) | 0.781 (0.68 - 0.88) | 0.809 (0.69 - 0.93) |
| p-tau181/$A\beta_{1-42}$ | | 0.88 (0.83 - 0.93)*** | 0.898 (0.83 - 0.96)*** | 0.836 (0.77 - 0.90)* | 0.861 (0.77 - 0.95) | 0.931 (0.86 - 1)* |
| p-tau231/$A\beta_{1-42}$ | | 0.781 (0.72 - 0.84)* | 0.771 (0.68 - 0.86) | 0.763 (0.68 - 0.84) | 0.8 (0.7 - 0.9) | 0.827 (0.71 - 0.94) |
| N ($A\beta^-/A\beta^+$) | @ CL20 | 142/91 | 62/38 | 124/44 | 56/22 | 18/47 |
| Base model | | 0.736 (0.67 - 0.8) | 0.739 (0.64 - 0.84) | 0.705 (0.61 - 0.8) | 0.782 (0.67 - 0.89) | 0.775 (0.66 - 0.9) |
| $A\beta_{1-42}$ | | 0.782 (0.72 - 0.84)* | 0.834 (0.76 - 0.91)* | 0.752 (0.67 - 0.84) | 0.853 (0.76 - 0.94) | 0.874 (0.77 - 0.98)* |
| $A\beta_{1-42/1-40}$ | | 0.793 (0.73 - 0.85)* | 0.812 (0.73 - 0.90)* | 0.751 (0.67 - 0.84) | 0.834 (0.74 - 0.93) | 0.901 (0.81 - 0.99)* |
| p-tau181 | | 0.870 (0.82 - 0.92)*** | 0.914 (0.85 - 0.97)** | 0.823 (0.75 - 0.90)* | 0.877 (0.79 - 0.97)* | 0.898 (0.81 - 0.99)* |
| p-tau231 | | 0.753 (0.69 - 0.82) | 0.771 (0.68 - 0.87) | 0.727 (0.64 - 0.82) | 0.8 (0.69 - 0.91) | 0.816 (0.71 - 0.92) |
| p-tau181/$A\beta_{1-42}$ | | 0.889 (0.84 - 0.94)*** | 0.952 (0.9 - 1)*** | 0.843 (0.77 - 0.92)** | 0.92 (0.84 - 1)* | 0.916 (0.83 - 1)* |
| p-tau231/$A\beta_{1-42}$ | | 0.776 (0.71 - 0.84)* | 0.798 (0.71 - 0.88) | 0.752 (0.67 - 0.84) | 0.825 (0.73 - 0.92) | 0.848 (0.76 - 0.94) |
| N ($A\beta^-/A\beta^+$) | @ CL25 | 146/87 | 66/34 | 128/40 | 60/18 | 18/47 |
| Base model | | 0.737 (0.67 - 0.81) | 0.742 (0.64 - 0.84) | 0.718 (0.62 - 0.81) | 0.806 (0.7 - 0.91) | 0.775 (0.66 - 0.9) |
| $A\beta_{1-42}$ | | 0.781 (0.72 - 0.84)* | 0.812 (0.73 - 0.89) | 0.756 (0.67 - 0.84) | 0.856 (0.76 - 0.95) | 0.874 (0.77 - 0.98)* |
| $A\beta_{1-42/1-40}$ | | 0.798 (0.74 - 0.86)* | 0.81 (0.72 - 0.9) | 0.768 (0.69 - 0.85)* | 0.857 (0.77 - 0.95) | 0.901 (0.81 - 0.99)* |
| p-tau181 | | 0.89 (0.84 - 0.94)*** | 0.956 (0.92 - 0.99) | 0.859 (0.79 - 0.93)** | 0.947 (0.9 - 0.99) | 0.898 (0.81 - 0.99)* |
| p-tau231 | | 0.754 (0.69 - 0.82) | 0.783 (0.69 - 0.88) | 0.728 (0.63 - 0.82) | 0.834 (0.73 - 0.94) | 0.816 (0.71 - 0.92) |

| | | | | | |
|---|---|---|---|---|---|
| p-tau181/A$\beta_{1-42}$ | 0.909 (0.87 - 0.95)*** | 0.97 (0.94 - 1) | 0.875 (0.8 - 0.94)** | 0.963 (0.93 - 1) | 0.916 (0.83 - 1)* |
| p-tau231/A$\beta_{1-42}$ | 0.775 (0.71 - 0.84)* | 0.813 (0.72 - 0.9) | 0.751 (0.66 - 0.84) | 0.863 (0.76 - 0.97) | 0.848 (0.76 - 0.94) |

Adjusted models were not performed within the MCI sub group for assessment 2 given the small sample sizes at CL15, N Aβ- = 17, N Aβ+ = 5, CL20 & 25, N Aβ- = 16, N Aβ+ = 6. P-values from the comparison between the base model (age, gender, tracer & *APOE* ε4) and the base model with the biomarker. * P<0.05, **P<0.001, ***P<0.0001

Supplementary Table 8: Positive and negative predictive values from ROC tests using the plasma p-tau181/A$\beta_{1-42}$ ratio to predict A$\beta$-PET status at three CL thresholds, two assessments and for individual and model-based biomarkers.

| Unadjusted/individual biomarker | | | | | | |
|---|---|---|---|---|---|---|
| CL threshold | Assessment | | AUC (95%CI) | Threshold | PPV | NPV |
| 15 | 1 | WC | 0.859 (0.81 - 0.91) | 1.277 | 86.96 | 82.27 |
| 20 | 1 | | 0.883 (0.83 - 0.93) | 1.483 | 88.89 | 87.5 |
| 25 | 1 | | 0.905 (0.86 - 0.95) | 1.483 | 88.89 | 90.13 |
| 15 | 2 | | 0.883 (0.81 - 0.95) | 1.139 | 87.5 | 85 |
| 20 | 2 | | 0.942 (0.89 - 1) | 1.139 | 85 | 93.33 |
| 25 | 2 | | 0.97 (0.94 - 1) | 1.28 | 91.18 | 95.45 |
| 15 | 1 | CN | 0.798 (0.72 - 0.88) | 1.292 | 81.4 | 84 |
| 20 | 1 | | 0.832 (0.75 - 0.91) | 1.312 | 76.19 | 90.48 |
| 25 | 1 | | 0.873 (0.8 - 0.94) | 1.312 | 73.81 | 92.86 |
| 15 | 2 | | 0.843 (0.74 - 0.94) | 1.139 | 82.61 | 85.45 |
| 20 | 2 | | 0.912 (0.83 - 1) | 1.079 | 73.08 | 94.23 |
| 25 | 2 | | 0.958 (0.92 - 1) | 1.139 | 73.91 | 98.18 |
| 15 | 1 | CI | 0.892 (0.78 - 1) | 1.006 | 94 | 80 |
| 20 | 1 | | 0.908 (0.82 - 1) | 1.478 | 95.56 | 80 |
| 25 | 1 | | 0.908 (0.82 - 1) | 1.478 | 95.56 | 80 |
| 15 | 2 | | 0.965 (0.89 - 1) | 1.277 | 100 | 83.33 |
| 20 | 2 | | 1 (1 - 1) | 1.277 | 100 | 100 |
| 25 | 2 | | 1 (1 - 1) | 1.277 | 100 | 100 |
| ROC values from models including biomarker, age, gender, tracer and *APOE* ε4 allele status | | | | | | |
| CL threshold | Assessment | | AUC (95%CI) | Threshold | PPV | NPV |
| 15 | 1 | WC | 0.88 (0.83 - 0.93) | 1.277 | 87.21 | 79.59 |
| 20 | 1 | | 0.889 (0.84 - 0.94) | 1.483 | 86.9 | 87.92 |
| 25 | 1 | | 0.909 (0.87 - 0.95) | 1.483 | 83.52 | 92.25 |
| 15 | 2 | | 0.898 (0.83 - 0.96) | 1.139 | 94.29 | 83.08 |
| 20 | 2 | | 0.952 (0.9 - 1) | 1.139 | 87.5 | 95 |
| 25 | 2 | | 0.97 (0.94 - 1) | 1.28 | 91.18 | 95.45 |
| 15 | 1 | CN | 0.836 (0.77 - 0.9) | 1.292 | 56.47 | 91.57 |
| 20 | 1 | | 0.843 (0.77 - 0.92) | 1.312 | 77.5 | 89.84 |
| 25 | 1 | | 0.875 (0.8 - 0.94) | 1.312 | 72.73 | 93.55 |
| 15 | 2 | | 0.861 (0.77 - 0.95) | 1.139 | 76.92 | 86.54 |
| 20 | 2 | | 0.92 (0.84 - 1) | 1.079 | 72.41 | 97.96 |
| 25 | 2 | | 0.963 (0.93 - 1) | 1.139 | 68 | 98.11 |
| 15 | 1 | CI | 0.931 (0.86 - 1) | 1.006 | 95.65 | 68.42 |
| 20 | 1 | | 0.916 (0.83 - 1) | 1.478 | 93.75 | 88.24 |
| 25 | 1 | | 0.916 (0.83 - 1) | 1.478 | 93.75 | 88.24 |

Across both the whole cohort and the MCI groups, both PPV and NPV increased with increasing CL thresholds, whilst for the CN group it decreased from 81.4% at the 15CL threshold, down to 76.2% and

73.8% at the 20 and 25 CL thresholds. Optimal PPV within the CN group was 92.86% at the 25CL threshold, while optimal NPV for the MCI group was 95.56% also at the 25CL threshold.

Supplementary Table 9: Multivariate ROC modelling for plasma markers to predict Aβ-PET

| Cohort | Visit | Threshold (CL) | Biomarker combination | AUC (95% CI) | p-value |
|---|---|---|---|---|---|
| WC | 1 | 15 | p-tau181/ $A\beta_{1-42}$ ratio | 0.88 (0.83 - 0.93) | |
| | 1 | | $A\beta_{1-42}$ + p-tau231 + p-tau181 | 0.879 (0.83 - 0.93) | 0.595 |
| | 1 | 20 | p-tau181/$A\beta_{1-42}$ ratio | 0.889 (0.84 - 0.94) | |
| | 1 | | $A\beta_{1-42/1-40}$ + $A\beta_{1-40}$ + p-tau181 | 0.892 (0.85 - 0.94) | 0.426 |
| | 1 | 25 | p-tau181/$A\beta_{1-42}$ ratio | 0.909 (0.87 - 0.95) | |
| | 1 | | p-tau181 + $A\beta_{1-42}$ | 0.909 (0.87 - 0.95) | 0.81 |
| | 2 | 15 | p-tau181/$A\beta_{1-42}$ ratio | 0.898 (0.83 - 0.96) | |
| | 2 | | $A\beta_{1-42}$ + p-tau231 + p-tau181 | 0.909 (0.84 - 0.97) | 0.287 |
| | 2 | 20 | p-tau181/$A\beta_{1-42}$ ratio | 0.952 (0.9 - 1) | |
| | 2 | | $A\beta_{1-42/1-40}$ + $A\beta_{1-40}$ + p-tau181 | 0.96 (0.92 - 1) | 0.334 |
| | 2 | 25 | p-tau181/$A\beta_{1-42}$ ratio | 0.97 (0.94 - 1) | |
| | 2 | | p-tau181 + $A\beta_{1-42}$ | 0.971 (0.94 - 1) | 0.697 |
| CN | 1 | 15 | p-tau181/$A\beta_{1-42}$ ratio | 0.836 (0.77 - 0.9) | |
| | 1 | | p-tau181 + $A\beta_{1-42}$ | 0.834 (0.77 - 0.9) | 0.166 |
| | 1 | 20 | p-tau181/$A\beta_{1-42}$ ratio | 0.843 (0.77 - 0.92) | |
| | 1 | | p-tau181 + $A\beta_{1-42}$ | 0.84 (0.77 - 0.91) | 0.492 |
| | 1 | 25 | p-tau181/$A\beta_{1-42}$ ratio | 0.875 (0.8 - 0.94) | |
| | 1 | | p-tau181 + $A\beta_{1-42}$ | 0.876 (0.81 - 0.94) | 0.44 |
| | 2 | 15 | p-tau181/AB42 ratio | 0.861 (0.77 - 0.95) | |
| | 2 | | p-tau181 + $A\beta_{1-42}$ | 0.869 (0.78 - 0.96) | 0.479 |
| | 2 | 20 | p-tau181/$A\beta_{1-42}$ ratio | 0.92 (0.84 - 1) | |
| | 2 | | p-tau181 + $A\beta_{1-42}$ | 0.93 (0.85 - 1) | 0.422 |
| | 2 | 25 | p-tau181/$A\beta_{1-42}$ ratio | 0.963 (0.93 - 1) | |
| | 2 | | p-tau181 + $A\beta_{1-42}$ | 0.962 (0.92 - 1) | 0.862 |
| CI | 1 | 15 | p-tau181/$A\beta_{1-42}$ ratio | 0.931 (0.86 - 1) | |
| | 1 | | $A\beta_{1-42/1-40}$ + p-tau181 | 0.984 (0.96 - 1) | 0.0876 |
| | 1 | 20 | p-tau181/$A\beta_{1-42}$ ratio | 0.916 (0.83 - 1) | |
| | 1 | | $A\beta_{1-42/1-40}$ + p-tau181 | 0.949 (0.9 - 1) | 0.279 |
| | 1 | 25 | p-tau181/$A\beta_{1-42}$ ratio | 0.916 (0.83 - 1) | |
| | 1 | | $A\beta_{1-42/1-40}$ + p-tau181 | 0.949 (0.9 - 1) | 0.279 |
| | 2 | 15 | - | - | - |
| | 2 | | - | - | - |
| | 2 | 20 | - | - | - |
| | 2 | | - | - | - |
| | 2 | 25 | - | - | - |
| | 2 | | - | - | - |

***Multivariate panel is no better than p-tau181/A$\beta_{1-42}$***

Given the best performing marker was the p-tau181/A$\beta_{1-42}$ ratio, we compared whether a multivariate model using multiple individual markers would out-perform the single ratio. Combining all possible plasma measures (individual markers and ratio's) into one model using the stepAIC function, and doing this at each of the three CL thresholds, in the complete, CN and MCI groups, it was clear that combining multiple markers into a model performed no better to predict Aβ-PET compared with the model including the p-tau181/A$\beta_{1-42}$ ratio alone. Further results from the multivariate comparisons are shown in Supplementary Table 8.

Supplementary Table 10: Plasma markers to predict CSF $A\beta_{1-42}$ levels (split at <1054 ==positive)

| Biomarker | Complete sample AUC (95%CI) | CU AUC (95%CI) | CI AUC (95%CI) |
|---|---|---|---|
| N ($A\beta^-/A\beta^+$) | 79/76 | 70/36 | 9/40 |
| $A\beta_{1-42}$ | 0.665 (0.58 - 0.75) | 0.697 (0.59 - 0.8) | 0.608 (0.37 - 0.85) |
| $A\beta_{1-42/1-40}$ | 0.695 (0.61 - 0.78) | 0.732 (0.63 - 0.83) | 0.658 (0.44 - 0.88) |
| $A\beta_{1-40}$ | 0.515 (0.42 - 0.61) | 0.513 (0.4 - 0.63) | 0.481 (0.26 - 0.7) |
| p-tau231 | 0.601 (0.51 - 0.69) | 0.587 (0.47 - 0.7) | 0.664 (0.46 - 0.87) |
| p-tau181 | 0.8 (0.73 - 0.88) | 0.755 (0.65 - 0.86) | 0.881 (0.78 - 0.98) |
| p-tau181/$A\beta_{1-42}$ | 0.816 (0.74 - 0.89) | 0.786 (0.69 - 0.88) | 0.858 (0.75 - 0.97) |
| p-tau231/$A\beta_{1-42}$ | 0.648 (0.56 - 0.73) | 0.654 (0.54 - 0.77) | 0.661 (0.47 - 0.85) |

Supplementary Table 11: QC panel tested in $A\beta_{1-40}$ and $A\beta_{1-42}$ over 16 test runs. There was a single re-run for $A\beta_{1-40}$ and $A\beta_{1-42}$ while 2 re-runs were needed for the p-tau assays.

| | **$A\beta_{1-40}$ (pg/mL)** | | **$A\beta_{1-42}$ (pg/mL)** | |
|---|---|---|---|---|
| **Test run** | **QC1** | **QC2** | **QC1** | **QC2** |
| T210583 | 5,51 | 7,20 | 3,87 | 4,84 |
| T210591 | 5,53 | 7,22 | 3,11 | 5,13 |
| T210595 | 5,58 | na | 3,97 | na |
| T210603 | 5,88 | 8,04 | 3,86 | 5,74 |
| T210610 | 5,30 | 6,91 | 4,04 | 5,44 |
| T210614 | 5,84 | 7,45 | 3,71 | 5,45 |
| T210618 | 5,43 | 6,89 | 3,90 | 5,44 |
| T210622 | 5,71 | 7,47 | 3,90 | 5,98 |
| T210627 | 5,28 | 7,49 | 3,84 | 6,00 |
| T210628 | 5,54 | 7,23 | 4,26 | 5,97 |
| T210633 | 4,87 | 7,47 | 4,13 | 6,47 |
| T210645 | 5,79 | 8,15 | 4,01 | 3,37 |
| T210648 | 5,81 | 7,53 | 3,80 | 6,35 |
| T210654 | 4,96 | 6,71 | 3,92 | 5,37 |
| T210659 | 5,21 | 7,87 | 3,81 | 5,94 |
| T210676 | 5,89 | 7,86 | 3,38 | 6,02 |
| **Mean (pg/mL)** | **5,51** | **7,43** | **3,84** | **5,57** |
| **Variation (CV%)** | **5,8** | **5,7** | **7,1** | **13,5** |

Supplementary Table 12: Inter-run variation on the AEB values of the calibrator points (CAL1-CAL7) for the $A\beta_{1-40}$, $A\beta_{1-42}$, p-tau181 tau and p-tau231 assay. The calibrator concentrations are described, the mean AEB signal given by the instrument and the inter-run variation in %CV calculated on AEB values.

| Calibrator point | $A\beta_{1-40}$ (pg/mL) | Mean AEB | Inter-run CV% |
|---|---|---|---|
| CAL 1 | 20 | 7,071 | 8,4 |
| CAL 2 | 10 | 2,479 | 7,8 |
| CAL 3 | 8 | 1,744 | 11,1 |
| CAL 4 | 6 | 1,088 | 7,8 |
| CAL 5 | 4 | 0,545 | 8,9 |
| CAL 6 | 2 | 0,157 | 7,9 |
| CAL 7 | 1 | 0,052 | 10,3 |

| Calibrator point | $A\beta_{1-42}$ (pg/mL) | Mean AEB | Inter-run CV% |
|---|---|---|---|
| CAL 1 | 18,4 | 2,173 | 9,3 |
| CAL 2 | 10,4 | 0,887 | 8,0 |
| CAL 3 | 7,0 | 0,432 | 9,0 |
| CAL 4 | 4,9 | 0,260 | 8,9 |
| CAL 5 | 2,6 | 0,080 | 8,1 |
| CAL 6 | 1,4 | 0,028 | 12,1 |
| CAL 7 | 1,0 | 0,021 | 16,4 |

| Calibrator point | p-tau181 (pg/mL) | Mean AEB | Inter-run CV% |
|---|---|---|---|
| CAL 1 | 50 | 0,808 | 9,2 |
| CAL 2 | 25 | 0,324 | 9,3 |
| CAL 3 | 12,5 | 0,146 | 9,6 |
| CAL 4 | 6,25 | 0,074 | 11,4 |
| CAL 5 | 3,125 | 0,046 | 8,2 |
| CAL 6 | 1,5625 | 0,032 | 6,9 |
| CAL 7 | 0,78125 | 0,025 | 8,3 |

| Calibrator point | p-tau231 (pg/mL) | Mean AEB | Inter-run CV% |
|---|---|---|---|
| CAL 1 | 50 | 1,309 | 10,0 |
| CAL 2 | 12,5 | 0,277 | 10,8 |
| CAL 3 | 6,25 | 0,143 | 18,8 |
| CAL 4 | 3,125 | 0,076 | 11,7 |
| CAL 5 | 1,5625 | 0,045 | 16,4 |
| CAL 6 | 0,78125 | 0,030 | 15,0 |
| CAL 7 | 0,390625 | 0,022 | 14,1 |

Supplementary Table 13: Summary table for the $A\beta_{1-40}$, $A\beta_{1-42}$, p-tau181 and p-tau231 assay: number of samples in the AIBL cohort testing below the measuring range (below Lower Limit Of Quantification or LLOQ), number of samples with a single value and number of samples where the variation on the duplicate testing was higher than 20%CV. The analytical LLOQ for $A\beta_{1-40}$ and $A\beta_{1-42}$ was respectively 1 pg/mL and 0.98 pg/mL based upon calibrator precision profile while the analytical LLOQ for p-tau181 and p-tau231 was respectively 0.81 and 0.38 pg/mL based upon calibrator precision profile.

| | $A\beta_{1-40}$ | $A\beta_{1-42}$ | p-tau181 | p-tau231 |
|---|---|---|---|---|
| #samples below LLOQ | 0 | 0 | 9 | 6 |
| #samples with single value | 6 | 27 | 15 | 9 |
| #samples with %CV>20 | 1 | 1 | 21 | 32 |

Supplementary Table 14: Overview of the accuracy data obtained with the calibrator points of the 4 Simoa assays. The three rows below each column show the mean back calculated concentration (BCC), the variation on the BCC (CV%) and the relative BCC as calculated against the theoretical value (BCC %).

| | **A$\beta_{1-40}$ calibrator point (pg/mL)** | | | | | | |
|---|---|---|---|---|---|---|---|
| Run ID | 20 | 10 | 8 | 6 | 4 | 2 | 1 |
| T210583 | 20,00 | 9,92 | 8,34 | 5,93 | 3,97 | 1,91 | 1,14 |
| T210591 | 20,01 | 9,78 | 8,24 | 6,10 | 3,95 | 1,84 | 1,06 |
| T210595 | 20,00 | 9,98 | 8,06 | 5,95 | 4,02 | 1,99 | 1,02 |
| T210603 | 20,00 | 9,98 | 8,09 | 5,86 | 4,12 | 1,92 | 1,03 |
| T210610 | 20,00 | 9,92 | 8,20 | 5,82 | 4,04 | 2,09 | na |
| T210614 | 20,00 | 9,97 | 8,06 | 5,98 | 3,98 | 1,92 | 1,10 |
| T210618 | 20,00 | 10,03 | 7,88 | 6,05 | 4,06 | 1,90 | 1,06 |
| T210622 | 20,00 | 10,02 | 7,98 | 5,97 | 4,07 | 2,01 | na |
| T210627 | 20,00 | 9,99 | 8,00 | 6,03 | 3,97 | 1,93 | 1,10 |
| T210628 | 20,00 | 10,10 | 7,57 | 6,43 | 4,01 | 1,89 | na |
| T210633 | 20,00 | 10,03 | 7,95 | 6,00 | 4,05 | 2,02 | na |
| T210645 | 20,00 | 10,07 | 7,85 | 6,08 | 4,15 | 1,88 | 1,05 |
| T210648 | 20,00 | 10,01 | 8,01 | 5,91 | 4,10 | 2,06 | na |
| T210654 | 20,00 | 10,17 | 7,63 | 6,15 | 4,18 | 2,01 | na |
| T210659 | 20,00 | 9,70 | 8,28 | 5,95 | 3,79 | 1,93 | 1,24 |
| T210676 | 20,01 | 10,04 | 7,51 | 6,58 | 4,23 | 1,80 | na |
| Mean | 20,00 | 9,98 | 7,98 | 6,05 | 4,04 | 1,94 | 1,09 |
| CV% | 0,02% | 1,16% | 3,07% | 3,30% | 2,59% | 4,05% | 6,14% |
| BCC (%) | 100,0 | 99,8 | 99,7 | 100,8 | 101,1 | 97,2 | 109,0 |

| | Aβ$_{1-42}$ calibrator point (pg/mL) | | | | | | |
|---|---|---|---|---|---|---|---|
| Run ID | 18,4 | 10,4 | 7,0 | 4,91 | 2,57 | 1,42 | 0,98 |
| T210583 | 18,40 | 10,46 | 6,75 | 5,20 | 2,56 | 1,25 | 0,93 |
| T210591 | 18,40 | 10,46 | 6,85 | 5,14 | 2,41 | 1,28 | 1,18 |
| T210595 | 18,40 | 10,43 | 6,88 | 5,06 | 2,57 | 1,34 | 0,94 |
| T210603 | 18,40 | 10,40 | 6,98 | 4,95 | 2,57 | 1,35 | 1,05 |
| T210610 | 18,42 | 10,18 | 7,23 | 5,18 | 2,52 | 1,12 | 0,41 |
| T210614 | 18,40 | 10,50 | 6,81 | 5,10 | 2,66 | 1,19 | 0,88 |
| T210618 | 18,40 | 10,43 | 6,87 | 5,08 | 2,51 | 1,40 | 0,93 |
| T210622 | 18,40 | 10,47 | 6,72 | 5,23 | 2,53 | 1,27 | 0,99 |
| T210627 | 18,40 | 10,44 | 6,83 | 5,15 | 2,49 | 1,13 | 1,25 |
| T210628 | 18,39 | 10,44 | 6,90 | 4,97 | 2,94 | 0,74 | na |
| T210633 | 18,40 | 10,48 | 6,36 | 5,26 | 2,68 | 1,15 | 0,55 |
| T210645 | 18,40 | 10,39 | 6,94 | 5,12 | 2,37 | 1,37 | 1,09 |
| T210648 | 18,39 | 10,46 | 6,75 | 5,20 | 2,57 | 1,23 | 0,98 |
| T210654 | 18,40 | 10,42 | 6,91 | 5,00 | 2,59 | 1,22 | 1,05 |
| T210659 | 18,40 | 10,47 | 6,85 | 5,12 | 2,47 | 1,27 | 1,12 |
| T210676 | 18,40 | 10,45 | 6,81 | 5,15 | 2,53 | 1,31 | 0,98 |
| Mean | 18,40 | 10,43 | 6,84 | 5,12 | 2,56 | 1,23 | 0,96 |
| CV% | 0,04% | 0,69% | 2,55% | 1,79% | 4,99% | 12,60% | 22,84% |
| BCC (%) | 100,0 | 100,3 | 97,7 | 104,3 | 99,6 | 86,4 | 97,5 |

| | p-tau181 calibrator point (pg/mL) | | | | | | |
|---|---|---|---|---|---|---|---|
| Run ID | 50 | 25 | 12,50 | 6,25 | 3,13 | 1,56 | 0,78 |
| T210583 | 52,21 | 24,54 | 11,86 | 6,25 | 3,20 | 1,71 | 0,75 |
| T210591 | 50,51 | 25,33 | 12,75 | 5,55 | 3,39 | 1,58 | 0,89 |
| T210595 | 51,36 | 25,01 | 12,03 | 5,99 | 3,37 | 1,89 | 1,04 |
| T210603 | 54,28 | 24,04 | 12,00 | 5,91 | 2,77 | 1,80 | 1,32 |
| T210610 | 53,90 | 24,49 | 12,14 | 5,72 | 2,68 | 2,06 | 1,13 |
| T210614 | 52,60 | 24,29 | 12,12 | 6,12 | 3,27 | 1,43 | 1,13 |
| T210618 | 56,30 | 24,14 | 10,60 | 5,87 | 3,23 | 1,90 | 1,19 |
| T210622 | 57,33 | 21,54 | 11,17 | 6,15 | 2,98 | 1,99 | 0,86 |
| T210627 | 54,74 | 23,48 | 11,50 | 5,99 | 3,57 | 1,59 | 0,88 |
| T210628 | 54,74 | 24,84 | 10,98 | 5,87 | 3,23 | 1,74 | 1,04 |
| T210633 | 53,80 | 23,96 | 12,43 | 5,05 | 3,31 | 2,46 | 1,81 |
| T210645 | nt | nt | nt | nt | nt | nt | nt |
| T210648 | 54,90 | 23,01 | 11,72 | 6,25 | 3,19 | 1,86 | 0,83 |
| T210654 | 52,31 | 24,51 | 12,07 | 5,98 | 3,31 | 1,79 | 0,92 |
| T210659 | 53,21 | 24,90 | 11,10 | 6,11 | 3,52 | 1,64 | 0,87 |
| T210676 | 51,83 | 24,64 | 12,29 | 5,69 | 3,78 | 1,56 | 0,70 |
| T210683 | 54,74 | 24,69 | 10,98 | 5,62 | 3,57 | 1,81 | 1,00 |
| Mean | 53,67 | 24,21 | 11,73 | 5,88 | 3,27 | 1,80 | 1,02 |
| CV% | 3,39% | 3,80% | 5,22% | 5,22% | 8,73% | 13,51% | 26,21% |
| BCC (%) | 107,3 | 96,8 | 93,8 | 94,1 | 104,7 | 115,2 | 130,9 |

| | p-tau231 calibrator point (pg/mL) | | | | | | |
|---|---|---|---|---|---|---|---|
| Run ID | 50 | 12,5 | 6,25 | 3,13 | 1,56 | 0,78 | 0,39 |
| T210583 | 56,73 | 11,62 | 5,22 | 2,87 | 1,64 | 1,02 | 0,36 |
| T210591 | 56,16 | 11,07 | 5,84 | 2,92 | 1,62 | 0,85 | 0,48 |
| T210595 | 53,02 | 11,89 | 5,72 | 3,24 | 1,61 | 0,84 | 0,39 |
| T210603 | 51,48 | 12,14 | 6,04 | 3,38 | 1,27 | 0,74 | 0,53 |
| T210610 | 54,50 | 10,60 | 6,68 | 3,12 | 1,28 | 0,84 | 0,59 |
| T210614 | 55,40 | 10,80 | 6,00 | 3,15 | 1,49 | 0,92 | 0,44 |
| T210618 | 54,70 | 12,30 | 5,89 | 2,53 | 1,58 | 0,89 | 0,54 |
| T210622 | 53,70 | 11,10 | 6,68 | 2,74 | 1,63 | 0,89 | 0,40 |
| T210627 | 57,60 | 11,70 | 4,68 | 2,97 | 1,82 | 0,83 | 0,63 |
| T210628 | 54,60 | 11,30 | 5,76 | 3,08 | 1,78 | 0,73 | 0,48 |
| T210633 | 52,60 | 13,50 | 5,28 | 2,80 | 1,64 | 0,78 | 0,54 |
| T210645 | 55,00 | 12,00 | 6,10 | 2,57 | 1,49 | 0,85 | 0,55 |
| T210648 | 49,90 | 12,10 | 7,39 | 2,70 | 1,43 | 0,79 | 0,43 |
| T210654 | 54,60 | 10,30 | 6,42 | 3,35 | 1,49 | 0,83 | 0,41 |
| T210659 | 52,80 | 12,50 | 5,72 | 2,94 | 1,50 | 1,04 | 0,39 |
| T210676 | 54,00 | 11,80 | 5,72 | 3,08 | 1,55 | 0,86 | 0,47 |
| T210683 | 53,70 | 12,00 | 5,38 | 3,24 | 1,65 | 0,82 | 0,47 |
| Mean | 54,15 | 11,69 | 5,91 | 2,98 | 1,56 | 0,85 | 0,48 |
| CV% | 3,48% | 6,67% | 10,78% | 8,62% | 9,43% | 9,74% | 16,25% |
| BCC (%) | 108,3 | 93,5 | 94,6 | 95,4 | 99,6 | 109,3 | 122,0 |

Supplementary Table 15: Intra-assay variation of the 4 Simoa assays. The intra-assay variation was calculated as %CV on the duplicate measurement of a sample if available. The total number of samples included in mentioned in the first line. Median and mean variation is reported as %CV.

| | $A\beta_{1-40}$ | $A\beta_{1-42}$ | p-tau181 | p-tau231 |
|---|---|---|---|---|
| Number of values | 381 | 360 | 372 | 380 |
| Minimum | 0 | 0 | 0 | 0 |
| 25% Percentile | 1,04 | 0,9925 | 2,9 | 3,2 |
| **Median** | **2,20** | **2,14** | **6,13** | **7,00** |
| 75% Percentile | 3,95 | 3,7 | 11,18 | 13,35 |
| Maximum | 25 | 27 | 50,1 | 74,5 |
| Mean | 3,2 | 2,7 | 7,9 | 9,3 |
| Std. Deviation | 3,403 | 2,578 | 6,595 | 8,477 |
| Std. Error of Mean | 0,1743 | 0,1359 | 0,3419 | 0,4349 |
| Lower 95% CI of mean | 2,865 | 2,457 | 7,197 | 8,444 |
| Upper 95% CI of mean | 3,551 | 2,991 | 8,542 | 10,15 |

Supplementary Figure 1: ROC curves for all biomarkers compared with the base model at Assessments 1 & 2 using a CL threshold at 20CL.

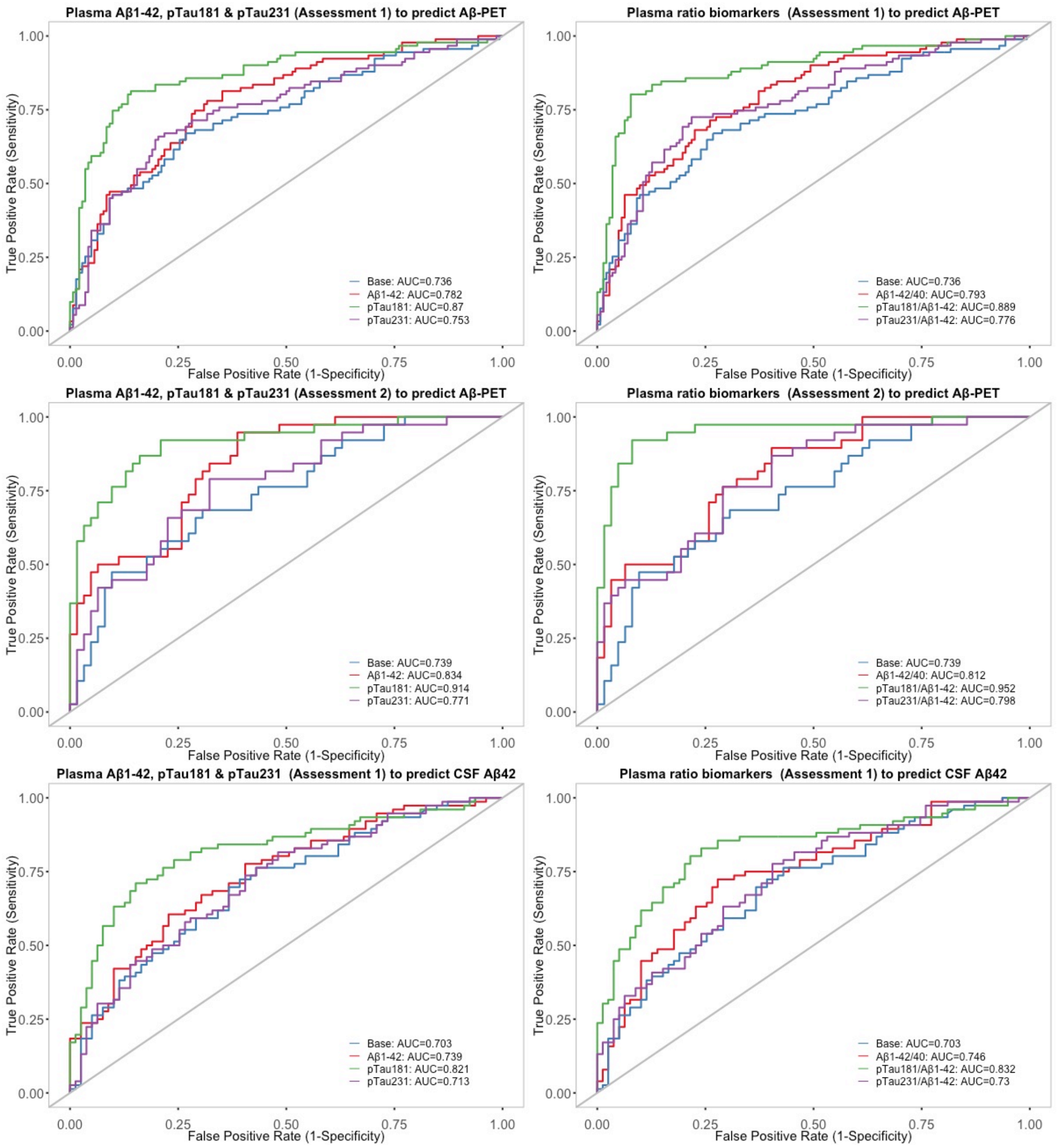

Supplementary Figure 2: Agreement between Aβ-PET and plasma p-tau181/$A\beta_{1-42}$ ratio at Assessment 2

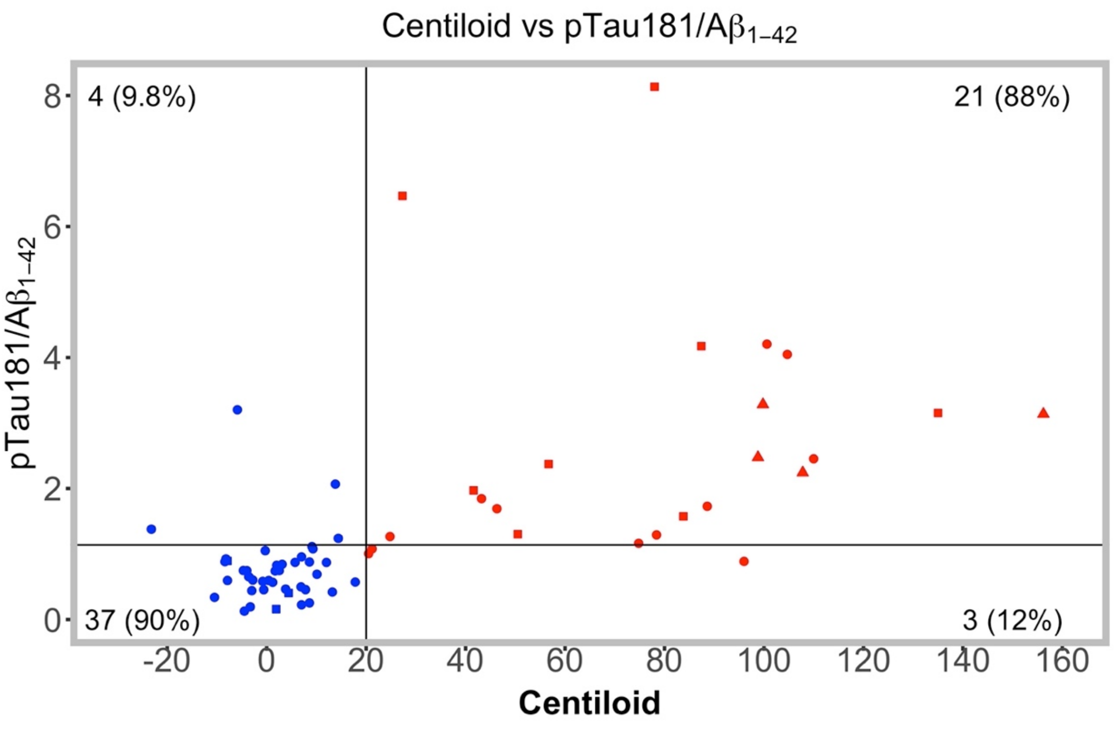

Threshold line for Aβ-PET was set at 20CL. Threshold for the p-tau181/$A\beta_{1-42}$ ratio was set using the Youden's Index (1.139) from the ROC model for Aβ-PET status with the CL threshold set at 20CL at Assessment 2. Red points represent participants who were Aβ-PET$^{+}$; blue points represent participants who were Aβ-PET$^{-}$. Circle points represent those participants who were CN, square points represent those participants with MCI, triangle points represent those participants with AD.

Supplementary Figure 3: Comparison of plasma vs CSF to predict Aβ-PET status

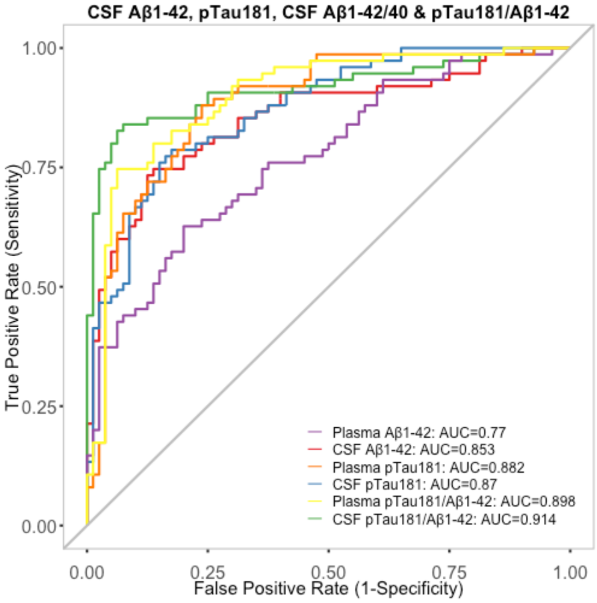

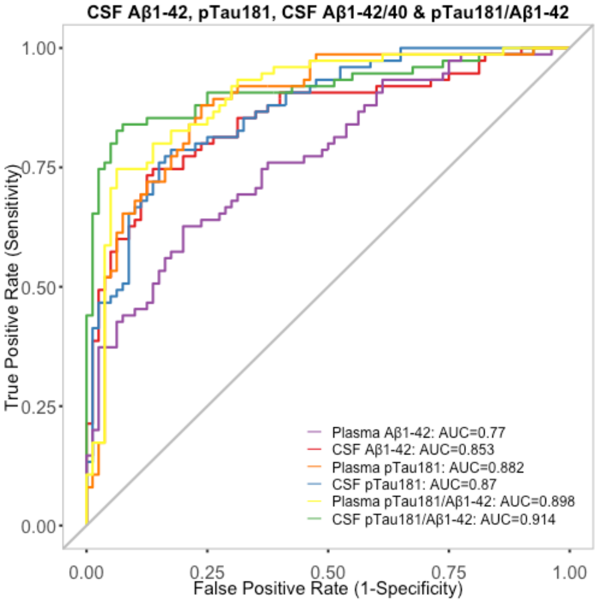

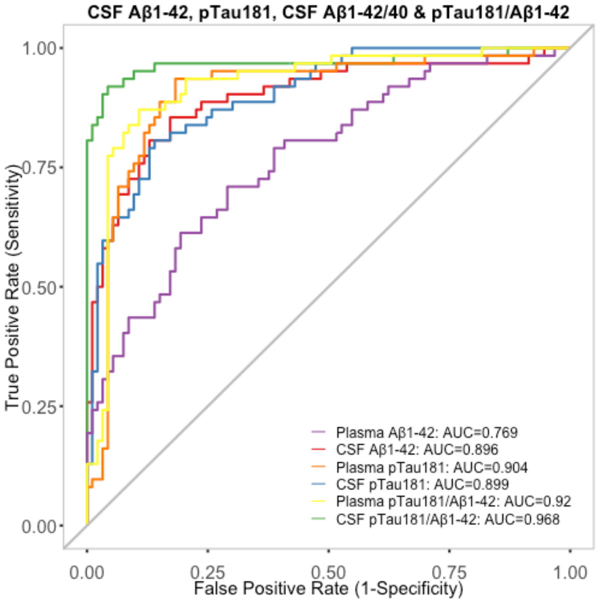

Supplementary Figure 4: Cognitive decline in CDR-SOB and the AIBL PACC score using the CSF between CN/CI groups and the p-tau181/A$\beta_{1\text{-}42}$ ratio as measured by linear mixed effects models

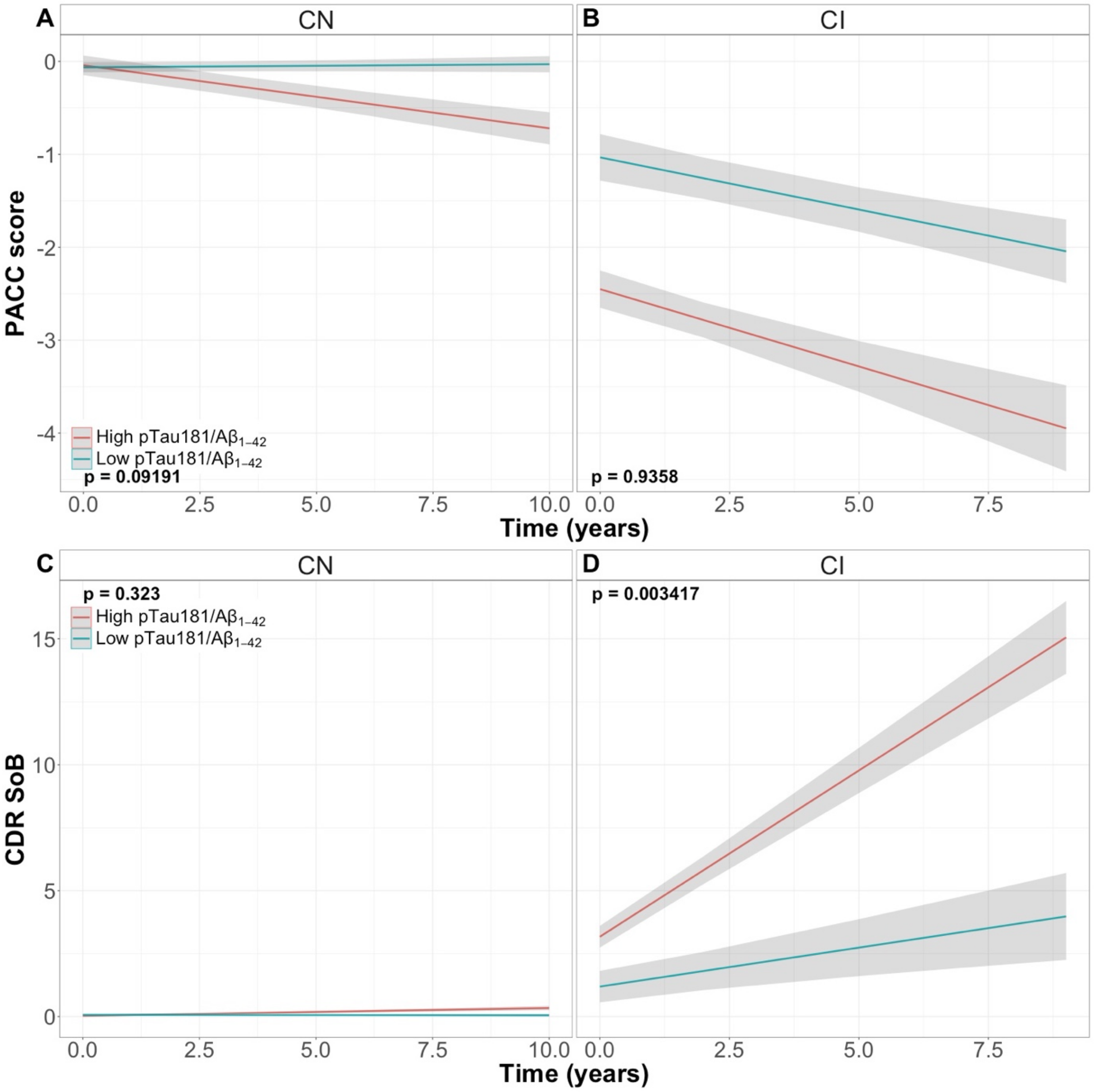

Time on the x-axis refers to the first AIBL collection whereby the plasma was collection. Sample sizes for cognitive collection points for the CSF sample set are shown in supplementary table 1. The binary p-tau181/A$\beta_{1\text{-}42}$ ratio was created using the Youden's Index (0.024) created from the ROC model using CSF p-tau181/A$\beta_{1\text{-}42}$ vs A$\beta$-PET using a CL threshold at 20CL.
